# Sleep in the Intensive Care Unit through the Lens of Breathing and Heart Rate Variability: A Cross-Sectional Study

**DOI:** 10.1101/2021.09.23.21264039

**Authors:** Wolfgang Ganglberger, Parimala Velpula Krishnamurthy, Syed A. Quadri, Ryan A. Tesh, Abigail A. Bucklin, Noor Adra, Madalena Da Silva Cardoso, Michael J. Leone, Aashritha Hemmige, Subapriya Rajan, Ezhil Panneerselvam, Luis Paixao, Jasmine Higgins, Muhammad Abubakar Ayub, Yu-Ping Shao, Brian Coughlin, Haoqi Sun, Elissa M. Ye, Sydney S. Cash, B. Taylor Thompson, Oluwaseun Akeju, David Kuller, Robert J. Thomas, M. Brandon Westover

## Abstract

**Background:** Full polysomnography, the gold standard of sleep measurement, is impractical for widespread use in the intensive care unit (ICU). Wrist-worn actigraphy and subjective sleep assessments do not measure sleep physiology adequately. Here, we explore the feasibility of estimating conventional sleep indices in the ICU with heart rate variability (HRV) and respiration signals using artificial intelligence methods.

**Methods:** We used deep learning models to stage sleep with HRV (through electrocardiogram) and respiratory effort (through a wearable belt) signals in critically ill adult patients admitted to surgical and medical ICUs, and in covariate-matched sleep laboratory patients. We analyzed the agreement of the determined sleep stages between the HRV- and breathing-based models, computed sleep indices, and quantified breathing variables during sleep.

**Results:** We studied 102 adult patients in the ICU across multiple days and nights, and 220 patients in a clinical sleep laboratory. We found that sleep stages predicted by HRV- and breathing-based models showed agreement in 60% of the ICU data and in 81% of the sleep laboratory data. In the ICU, deep NREM (N2 + N3) proportion of total sleep duration was reduced (ICU 39%, sleep laboratory 57%, p<0.01), REM proportion showed heavy-tailed distribution, and the number of wake transitions per hour of sleep (median = 3.6) was comparable to sleep laboratory patients with sleep-disordered breathing (median = 3.9). Sleep in the ICU was also fragmented, with 38% of sleep occurring during daytime hours. Finally, patients in the ICU showed faster and less variable breathing patterns compared to sleep laboratory patients.

**Conclusions:** Cardiovascular and respiratory signals encode sleep state information, which can be utilized to measure sleep state in the ICU. Using these easily measurable variables can provide automated information about sleep in the ICU.

## I. Introduction

Healthy sleep is a biological imperative(1, 2). Abnormal sleep impairs critical brain and body functions, including memory(3), learning, attention, and affective state(4–8), and regulation of blood pressure(9), inflammatory processes(10, 11), metabolic control(12–14), and stress responses(15–17). The intensive care unit (ICU) is associated with disrupted sleep, due to internal (e.g., pain, immunocompromisation, dyspnea and apnea) and external (e.g., noise, circadian mismatch) factors. Sleep can be so distorted in the ICU that conventional sleep stages become hard to recognize(18). Sleep disruption in the ICU contributes to delirium(19), difficult weaning from mechanical ventilation(20), and increased risk of autonomic, inflammatory and metabolic dysfunction(21).

Despite the urgency of improving sleep in the ICU, measuring sleep in this environment is challenging since conventional polysomnography is difficult to operationalize in the ICU setting(22). Subjective sleep estimates and movement analysis using actigraphy can provide crude assessments of sleep(22, 23), but are heavily confounded by common ICU experiences, including sedation, monitoring, illness, and immobility. No present method of measuring sleep in the ICU is satisfactory(24), thus alternative approaches are needed.

To address this need, our first aim for this study was to evaluate the validity of monitoring sleep physiology in ICU patients using easily obtainable biosignals, such as electrocardiogram (ECG) and respiration, using artificial intelligence methods. Although sleep states are commonly discerned through electroencephalogram (EEG) signals, they can also be decoded through analysis of non-EEG signals(25–27) since sleep modifies a variety of biosignals(28, 29), including blood pressure, heart rate, and respiration. Additionally, respiration and ECG measurements are easier to acquire, offer a more practical and repeatable diagnostic tool compared to EEG, and better reflect sleep physiology compared to actigraphy and subjective assessments. For this study, we used deep neural network models to determine sleep stages from heart rate variability (HRV), derived from ECG, and breathing signals, obtained with a single respiratory effort belt. We analyzed how well the HRV and breathing models agreed in determining sleep stages both in ICU patients and in age and sex-matched patients referred to a clinical sleep laboratory. We hypothesized that agreement between the two models signifies confidence in the determined sleep stage. To test this hypothesis, we used the sleep laboratory dataset to compare sleep staging performance between our model agreement method and the gold standard, which involves manual scoring of polysomnography EEG signals by experts. We also hypothesized that the HRV and breathing models would agree substantially in the ICU dataset, although we expected more disagreement compared to the sleep laboratory dataset given the extent of respiratory and cardiac physiology in critical illness. To characterize variables that could reduce the reliability of the presented HRV- and breathing-based sleep analysis in the ICU, we evaluated whether specific HRV- and respiratory features, medical conditions, severity of illness, and pharmacological drugs are associated with disagreement of these two sleep staging models.

The second aim of this study was to identify common patterns of sleep pathology in ICU patients compared to non-critically ill patients undergoing overnight diagnostic polysomnography recordings for suspected sleep disorders in the sleep laboratory. We computed common sleep statistics and respiratory variables such as respiratory rate and sleep breathing variability, for both cohorts, and tested the robustness of our results through sensitivity analysis.

Finally, sleep has been shown to be fragmented in the ICU(30), which prevents patients from getting adequate consolidated periods of rest. As such, the last aim of this study was to quantify sleep fragmentation in patients admitted to the ICU.

## II. Methods

### A. Study Oversight

Patients were enrolled after written consent in a randomized clinical trial, Investigation of Sleep in the Intensive Care Unit (NCT03355053, (31)), at the Massachusetts General Hospital (MGH) from June 2018 to November 2019. The clinical trial involved randomizing patients into three groups, where two of the groups received a low dose of dexmedetomidine (0.1 or 0.3 mcg/kg/h) overnight continuously for 11 hours, and the third group received placebo (normal saline). Exclusion criteria for the clinical trial include severe dementia, known pre-existing neurologic diseases or cognitive deficits, serious cardiac disease, severe liver dysfunction, severe renal dysfunction, and low likelihood of survival for 24 hours – all criteria can be found in the online supplement. The study was approved by the Mass General Brigham Institutional Review Board.

### B. Dataset – ICU Cohort

The sample size for this study was determined by starting with enrolled patients and then excluding all patients under 45 years of age and patients who had less than 2 hours of ECG or respiratory data. Patients were non-mechanically ventilated at the time of enrollment, although some were subsequently mechanically ventilated during the course of hospitalization; see **Table 1**. At the start of the trial, a respiratory belt (*Airgo*, a CE Class IIa certified wearable medical device (32), **Figure S1**) was placed around a patient’s chest, as close to the floating ribs as possible, until they were transferred outside of the ICU. The belt contains a conductive silver band that measures respiratory effort by sensing changes in electrical resistance that correspond to changes in belt length induced by thoracic movements. The amplitude values of the belt were not calibrated. The sampling frequency was 10 Hz. In addition to demographic data, we collected information regarding labs, medications, vital signs, and ICD-10 codes from the hospital’s electronic medical records. Vital signs at higher time resolution (0.5 Hz) and electrocardiogram (ECG) (256 Hz) data were collected from the bedside telemetry monitors over the hospital network using BedMaster software (Excel Medical, Jupiter, FL). Signals collected through bedside monitors and the wearable respiratory device both contained real-time timestamps; correct alignment was manually reviewed for all patients. Charlson Comorbidity Index(33) and Sequential Organ Failure Assessment (SOFA)(34) scores were computed.

**Table 1.**
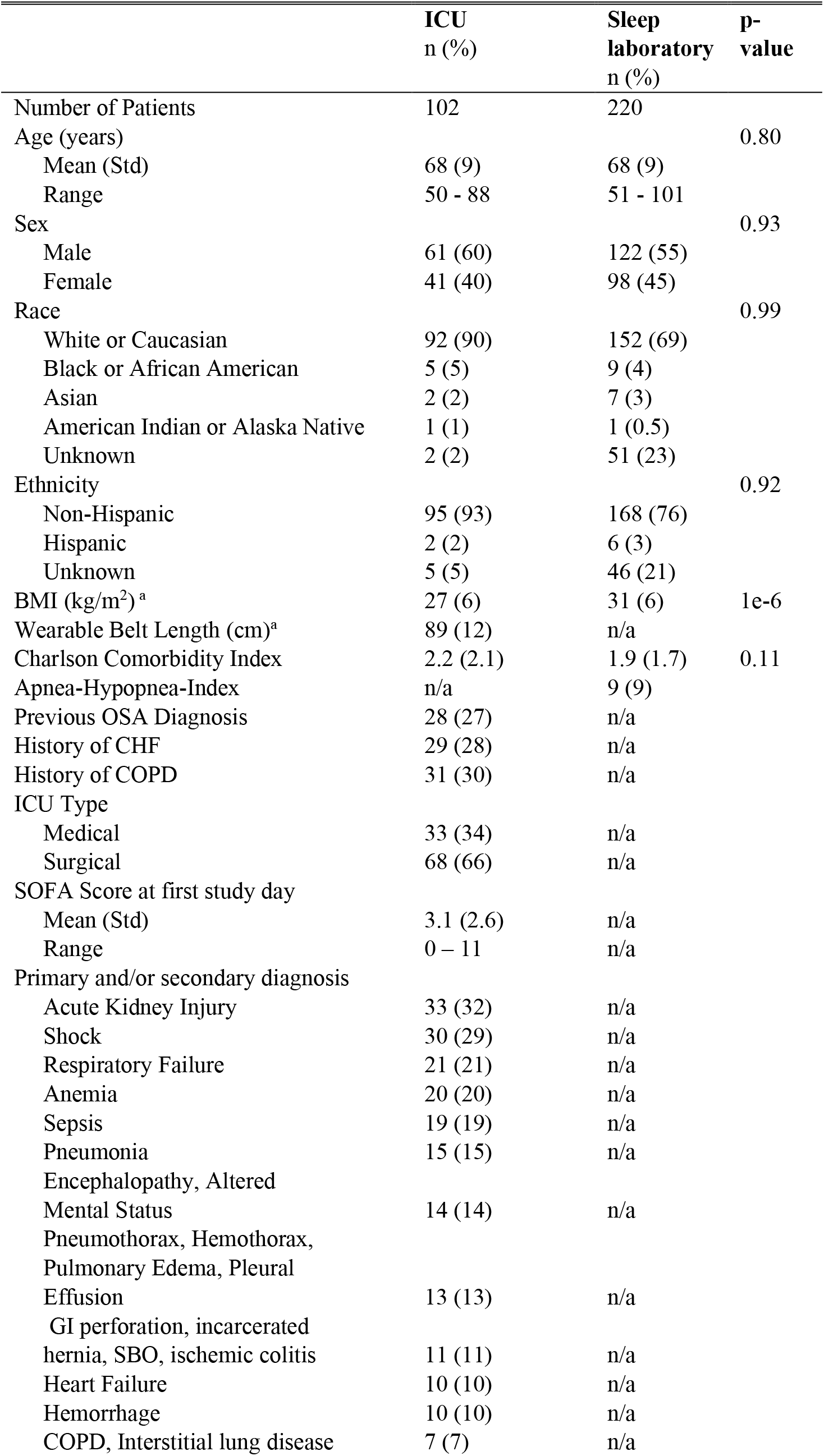
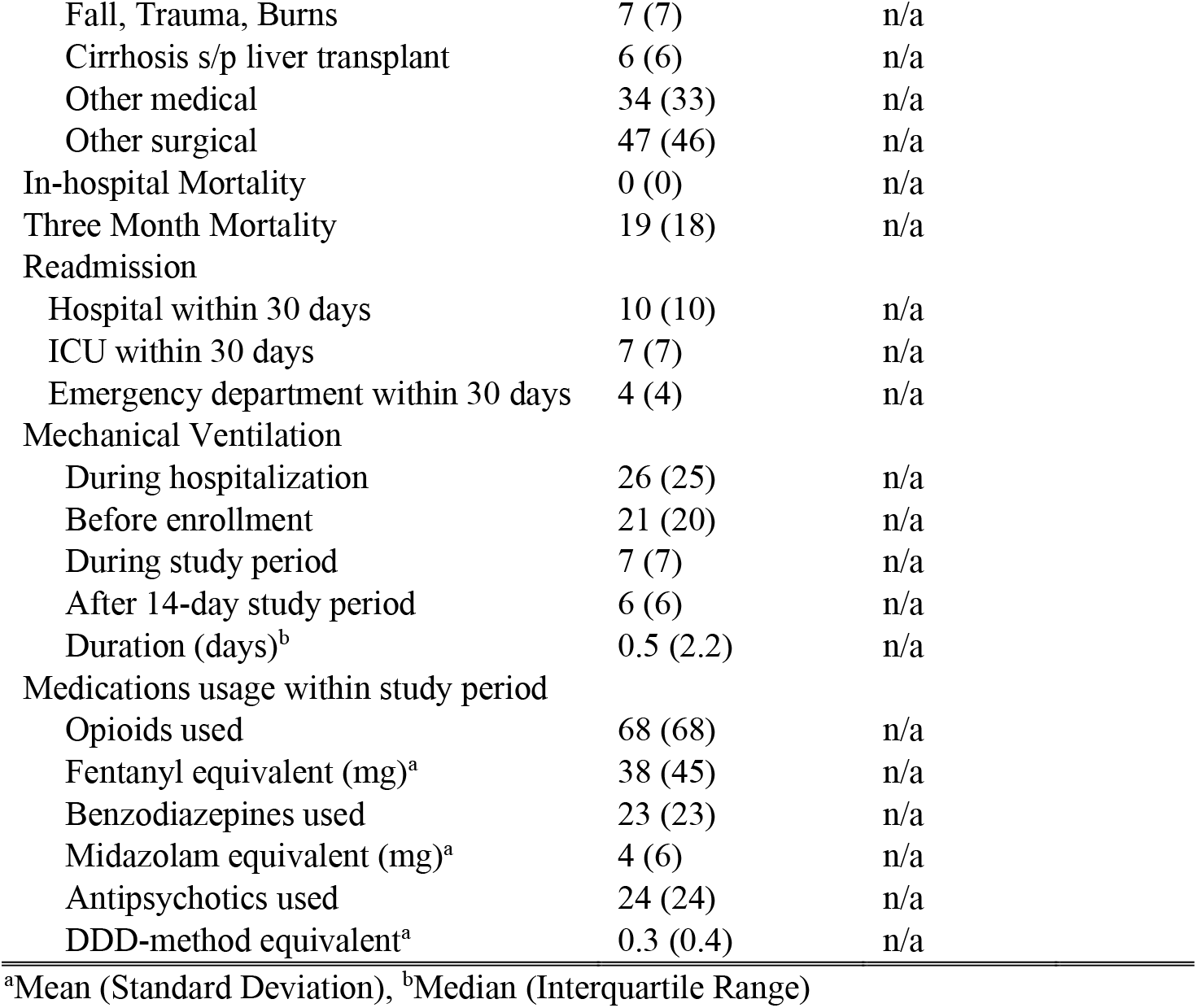
Baseline Characteristics

|  | ICU<br>n (%) | Sleep<br>laboratory<br>n (%) | p-value |
| --- | --- | --- | --- |
| Number of Patients | 102 | 220 |  |
| Age (years) |  |  | 0.80 |
| Mean (Std) | 68 (9) | 68 (9) |  |
| Range | 50 - 88 | 51 - 101 |  |
| Sex |  |  | 0.93 |
| Male | 61 (60) | 122 (55) |  |
| Female | 41 (40) | 98 (45) |  |
| Race |  |  | 0.99 |
| White or Caucasian | 92 (90) | 152 (69) |  |
| Black or African American | 5 (5) | 9 (4) |  |
| Asian | 2 (2) | 7 (3) |  |
| American Indian or Alaska Native | 1 (1) | 1 (0.5) |  |
| Unknown | 2 (2) | 51 (23) |  |
| Ethnicity |  |  | 0.92 |
| Non-Hispanic | 95 (93) | 168 (76) |  |
| Hispanic | 2 (2) | 6 (3) |  |
| Unknown | 5 (5) | 46 (21) |  |
| BMI (kg/m<sup>2</sup>)<sup>a</sup> | 27 (6) | 31 (6) | 1e-6 |
| Wearable Belt Length (cm)<sup>a</sup> | 89 (12) | n/a |  |
| Charlson Comorbidity Index | 2.2 (2.1) | 1.9 (1.7) | 0.11 |
| Apnea-Hypopnea-Index | n/a | 9 (9) |  |
| Previous OSA Diagnosis | 28 (27) | n/a |  |
| History of CHF | 29 (28) | n/a |  |
| History of COPD | 31 (30) | n/a |  |
| ICU Type |  |  |  |
| Medical | 33 (34) | n/a |  |
| Surgical | 68 (66) | n/a |  |
| SOFA Score at first study day |  |  |  |
| Mean (Std) | 3.1 (2.6) | n/a |  |
| Range | 0 – 11 | n/a |  |
| Primary and/or secondary diagnosis |  |  |  |
| Acute Kidney Injury | 33 (32) | n/a |  |
| Shock | 30 (29) | n/a |  |
| Respiratory Failure | 21 (21) | n/a |  |
| Anemia | 20 (20) | n/a |  |
| Sepsis | 19 (19) | n/a |  |
| Pneumonia | 15 (15) | n/a |  |
| Encephalopathy, Altered |  |  |  |
| Mental Status | 14 (14) | n/a |  |
| Pneumothorax, Hemothorax, |  |  |  |
| Pulmonary Edema, Pleural |  |  |  |
| Effusion | 13 (13) | n/a |  |
| GI perforation, incarcerated |  |  |  |
| hernia, SBO, ischemic colitis | 11 (11) | n/a |  |
| Heart Failure | 10 (10) | n/a |  |
| Hemorrhage | 10 (10) | n/a |  |
| COPD, Interstitial lung disease | 7 (7) | n/a |  |

|  |  |  |
| --- | --- | --- |
| Fall, Trauma, Burns | 7 (7) | n/a |
| Cirrhosis s/p liver transplant | 6 (6) | n/a |
| Other medical | 34 (33) | n/a |
| Other surgical | 47 (46) | n/a |
| In-hospital Mortality | 0 (0) | n/a |
| Three Month Mortality | 19 (18) | n/a |
| Readmission |  |  |
| Hospital within 30 days | 10 (10) | n/a |
| ICU within 30 days | 7 (7) | n/a |
| Emergency department within 30 days | 4 (4) | n/a |
| Mechanical Ventilation |  |  |
| During hospitalization | 26 (25) | n/a |
| Before enrollment | 21 (20) | n/a |
| During study period | 7 (7) | n/a |
| After 14-day study period | 6 (6) | n/a |
| Duration (days)<sup>b</sup> | 0.5 (2.2) | n/a |
| Medications usage within study period |  |  |
| Opioids used | 68 (68) | n/a |
| Fentanyl equivalent (mg)<sup>a</sup> | 38 (45) | n/a |
| Benzodiazepines used | 23 (23) | n/a |
| Midazolam equivalent (mg)<sup>a</sup> | 4 (6) | n/a |
| Antipsychotics used | 24 (24) | n/a |
| DDD-method equivalent<sup>a</sup> | 0.3 (0.4) | n/a |

<sup>a</sup>Mean (Standard Deviation), <sup>b</sup>Median (Interquartile Range)

*Sleep Laboratory Cohort*: 404 patients who underwent overnight polysomnography (PSG) in the Massachusetts General Hospital sleep laboratory between January 2019 and January 2020 wore the same respiratory belt that was used in the ICU cohort. Participants were enrolled through verbal consent shortly before onset of the PSG. There were no exclusion criteria and enrollment stopped after reaching a target sample size of 404 patients. This study was also approved by the Mass General Brigham Institutional Review Board. To use sleep laboratory patients as control subjects for comparison with ICU patients, we applied the same exclusion criteria, i.e., excluding all patients under 45 years of age. We also balanced the distribution of age and sex between the ICU and sleep lab cohorts by applying k-nearest-neighbor matching, as previously described(35). We further stratified sleep lab patients according to their Apnea-Hypopnea-Index (AHI) severity into no-disordered breathing (AHI < 5) and disordered breathing (AHI > 15) groups and applied the same matching method for each of these subgroups. Seven trained sleep technicians annotated PSG studies as part of routine clinical care according to American Academy of Sleep Medicine guidelines (36).

### C. Biosignals Preprocessing

Non-physiological data and data with low signal quality were removed from both the ECG and respiratory effort belt signal. For the respiratory signal, this was done with an algorithm checking for high and constant amplitude (belt not worn) and for absence of reliable breath detection (low signal quality). For each patient, we normalized the respiratory signal by subtracting the mean and dividing by the standard deviation calculated from the 1-99% quantile clipped signal. We used the open-source PhysioNet Cardiovascular Signal Toolbox(37) to filter the ECG signal, extract R peaks based on the Pan Tompkins algorithm(38), and obtain a signal quality measure. We provide parameter settings for the preprocessing steps in the online supplement.

### D. Sleep Staging

We used deep neural network models that use heart rate variability (a binary sequence with 1 for a detected R-peak in the ECG, 0 else) and respiratory effort signals as inputs to assign a sleep stage (Wake, R, N1, N2 and N3) to every 30-second epoch. We previously validated this approach on datasets from the MGH sleep laboratory and the Sleep Heart Health Study(27). Here, we used a model trained with signals from the wearable respiratory belt as input. For visual representation of the resulting sleep stages for all patients, swimmer plots were created.

### E. Breathing Features

From the respiratory belt’s signal, we computed four features using a moving window approach:

i. Respiratory rate (RR): number of breaths (inspiratory peaks) detected in 10 seconds (moving window) x 6.
ii. Inter-breath-interval (IBI): time (seconds) between two consecutive breaths.
iii. Ventilation coefficient of variation (ventilation CVar): we first computed a proxy of minute ventilation as the sum of positive amplitude changes (inspiration) over 10 seconds and scaled it to a reference of one minute (multiplied by 6), then computed the coefficient of variation over a 30-second window.
iv. Variability index: we computed the coefficient of variation of the IBIs over a 30-second window, and defined the variability index as: variability index = (ventilation CVar + IBI CVar)/2, i.e., the mean of the coefficient of variation computed from the breathing timing (IBI) and breathing amplitude (ventilation).

### F. Statistical Analysis – Sleep Staging

*Definition - concordant and discordant sleep*. For all analyses in this study, we only used data where both HRV and breathing data was simultaneously available. We applied both the HRV and breathing-based sleep staging models individually on all data. Each 30-second epoch in the data was assigned a sleep stage by the HRV- and breathing-based models, yielding two hypnograms. Because disagreement in sleep staging in human experts is common(39), and because stages between wake, N1, N2, and N3 form a continuum, we defined *concordance* regarding the stage assigned to a given 30-second epoch of sleep if the models agreed to within one stage, and *discordance* if they did not. Specifically, we defined an ordinal progression of sleep depth for NREM sleep: W < N1 < N2 < N3, such that e.g., if the HRV and breathing-based models assigned W and N1, respectively, this would be considered concordant; whereas assignments of W and N2 would be considered discordant. For REM sleep R, models were considered concordant only if both assigned a stage of R. For completeness, the full set of concordant and discordant stage assignments were:

*Concordance*: (W, W), (N1, N1), (N2, N2), (N3, N3), (R, R), (W, N1), (N1, N2), (N2, N3).

*Discordance*: (W, N2), (W, N3), (W, R), (N1, N3), (R, N1), (R, N2), (R, N3).

For the sleep lab polysomnography data, sleep staging agreements between the models and the experts were measured with confusion matrices and Cohen’s kappa(40) for both concordant and discordant data.

*Sleep indices*. We split each patient’s data into 24-hour segments (*full day*), starting and ending at 08:00, and further defined *day* as 08:00 – 20:00 and *night* as 20:00 – 08:00. For every 24-hour segment, we obtained the HRV-based and breathing-based hypnograms, and computed the following *sleep indices*:

1. Total sleep duration (in hours).
2. Concordant sleep duration (in hours).
3. Discordant sleep duration (in hours).
4. Proportion of discordant sleep from total sleep (in %).
5. Sleep fraction (%): sleep duration divided by amount of data available.
6. Stage R (%), Stage N1 (%), Stage N2 (%), Stage N3 (%), Stage N2+N3(%): Time spent in a specified sleep stage divided by sleep duration.
7. Sleep fragmentation index (SFI): Number of sleep stage transitions from (N2, N3, R) to (N1, W) divided by sleep duration.
8. Wake transitions / hour: Number of sleep stage transitions from (N1, N2, N3, R) to (W) divided by sleep duration.

We further computed mean sleep indices across the models:

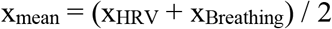

where x is any of the 8 sleep indices listed above.

To ensure robust conclusions regarding sleep indices, we performed sensitivity analysis, by computing sleep indices using three complementary approaches. The approaches vary by what segments are included for sleep index computation, and by which part of sleep (total sleep or concordant sleep) is used to compute the sleep indices:

A1. Inclusion criteria: Any amount of sleep. Sleep indices computed on total sleep.
A2. Inclusion criteria: At least two hours of concordant sleep. Sleep indices computed on total sleep.
A3. Inclusion criteria: At least two hours of concordant sleep. Sleep indices computed on concordant sleep.

For each approach, we carried out the following test procedure: For every subject we computed the mean sleep indices (mean x_mean_ across all 24-hour segments). We then applied a Mann-Whitney U (MWU) test (H0: ‘equal distribution for both groups’) and Mood’s median test (H0: ‘equal medians for both groups’) to assess statistical difference between the ICU and sleep lab (overall and AHI subgroups) subjects. We considered sleep indices to be significantly different for two groups (Mann-Whitney U) with a significantly different effect direction (Mood’s median test) if both tests resulted in a p-value of less than 0.05. We chose to apply non-parametric tests instead of t-tests because we found that none of the data were normally distributed in either the ICU or sleep lab group, where we define non-normally distributed to mean that either the Shapiro-Wilk or D’Agostino’s K-squared test rejects null hypothesis of normality for alpha=0.05. We report test results for all applied statistical tests.

*Sleep fragmentation in the ICU.* We characterized sleep fragmentation in the ICU(30) by the following metrics using HRV model-based sleep stages:

1. Proportion of day (08:00 – 20:00) spent asleep.
2. Proportion of night (20:00 – 08:00) spent asleep.
3. Proportion of sleep occurring in the day versus in the night.
4. Proportion of REM sleep occurring in the day versus in the night.
5. Number of sleep periods, with a duration of at least one minute, per 24 hours.
6. Number of sleep periods, with a duration of at least five minutes, per 24 hours.

*Subgroup Analysis.* The ICU patients were manually grouped according to their primary and main conditions, and the median sleep indices (HRV-based model, analysis approach A2) were computed for each group. The Kruskal-Wallis H test was applied to assess differences of individual sleep indices between groups.

*Latent feature representation of sleep.* To assess similarities and differences in the latent feature representation between sleep epochs in the ICU and the sleep lab, we computed unsupervised UMAPs (Uniform Manifold Approximation and Projection)(41) based on the sleep staging neural networks’ last hidden layer activations. UMAPs were created separately for the HRV-based model and breathing-based model across pooled sleep lab and ICU data, see supplement for details.

*Disagreement between HRV and breathing models - error analysis.* We hypothesized that the following variables affect discordance in the HRV- and breathing-based models: 1) specific features from cardiac and respiratory signals; 2) daily dosing of opioids, benzodiazepines, and antipsychotics; 3) SOFA score, a measure of a patient’s illness severity. HRV and breathing features of interest were: RR interval, RR root mean square of successive differences (RMSSD), HRV very low frequency power (VLF), HRV low frequency power (LF), HRV high frequency power (HF), inter-breath-intervals, respiratory rate, respiratory variability index, ventilation CVar, cardiopulmonary coupling(42) (CPC) low frequency coupling (LFC), CPC high frequency coupling (HFC). We computed these features for concordant and discordant sleep parts for each 24-hour segment and performed a Mann–Whitney U test with a significance level of 0.01 for each feature pair. Next, we computed features for each 24-hour segment and performed multilinear regression analysis with a LASSO penalty, with the features as independent variables and the discordant proportion (log-transformed) as the dependent variable. To test if daily administered doses of opioids (in Fentanyl milligram(43)), benzodiazepines (in Midazolam milligram(44, 45)), antipsychotics (DDD method(46)) and the SOFA score were associated with discordance, we computed Pearson and Spearman correlations between each of these variables and the proportion of discordant sleep for each 24-hour segment.

### G. Statistical Analysis – Breathing

For every 24-hour segment with at least two hours of concordant sleep, we computed the breathing features as described above from concordant sleep and for each sleep stage. For each patient we averaged the features across all nights. For ICU and sleep lab cohorts, we computed the mean and standard deviation of each feature and assessed statistically significant differences between cohorts with t-tests for Gaussian features or Mann-Whitney U tests and Mood’s median test (analogous to sleep stage analysis) for non-Gaussian features.

Results are reported in accordance with the Strengthening the Reporting of Observational Studies in Epidemiology (STROBE) guidelines(47). We provide the de-identified data, the analysis code and computational models used in this study on our GitHub page(48).

## III. Results

### B. Dataset

For the ICU cohort, 129 patients were enrolled in the clinical trial; we excluded two patients under 45 years old, and 25 patients for having less than 2 hours of ECG or respiratory data available from our analysis, resulting in a cohort size of 102 patients (41 females, 61 males). In the sleep lab cohort, out of 404 enrolled patients, 97 were excluded due to age under 45 years, resulting in a sample size of 307 before matching. Matching resulted in 220 (98 females, 122 males) sleep lab patients, 77 (40 females, 37 males) with an AHI < 5 and 52 (18 females, 34 males) with an AHI > 15. Age distributions were similar for ICU and matched groups, both for male and female subjects; see **Table S1** for details. **Table 1** summarizes the baseline characteristics of the ICU and the matched sleep lab cohort.

### C. Biosignals Preprocessing

After signal preprocessing, we obtained 6,728 hours (280 days) of ECG data, 3,886 hours (162 days) of breathing data, and 3,502 hours (146 days) of simultaneous ECG and breathing data for the ICU cohort. For the sleep lab cohort, the numbers were 1,634, 1,609, and 1,562 hours, respectively. Mean (SD) hours of data available per patient in the ICU (N=103) were ECG 66.0 (27.8), Breathing 38.1 (28.1), for simultaneous ECG and breathing signals 34.3 (24.6), and for the sleep lab (N=220) 7.4 (0.8), 7.3 (1.0), 7.1 (0.9) hours respectively.

### F. Statistical Analysis, Sleep Staging

In the sleep laboratory data, 131,157 out of 173,977 30-second epochs (75.4%) were assigned concordant sleep stages by the HRV- and breathing-based models. The models showed higher staging agreements with experts for concordant data than for discordant data, both when agreement was evaluated with Cohen’s kappa with AASM standard stages (W, R, N1, N2, N3), as well as with combined NREM stages (W, R, NREM), see **Figure 1**.

**Figure 1:**
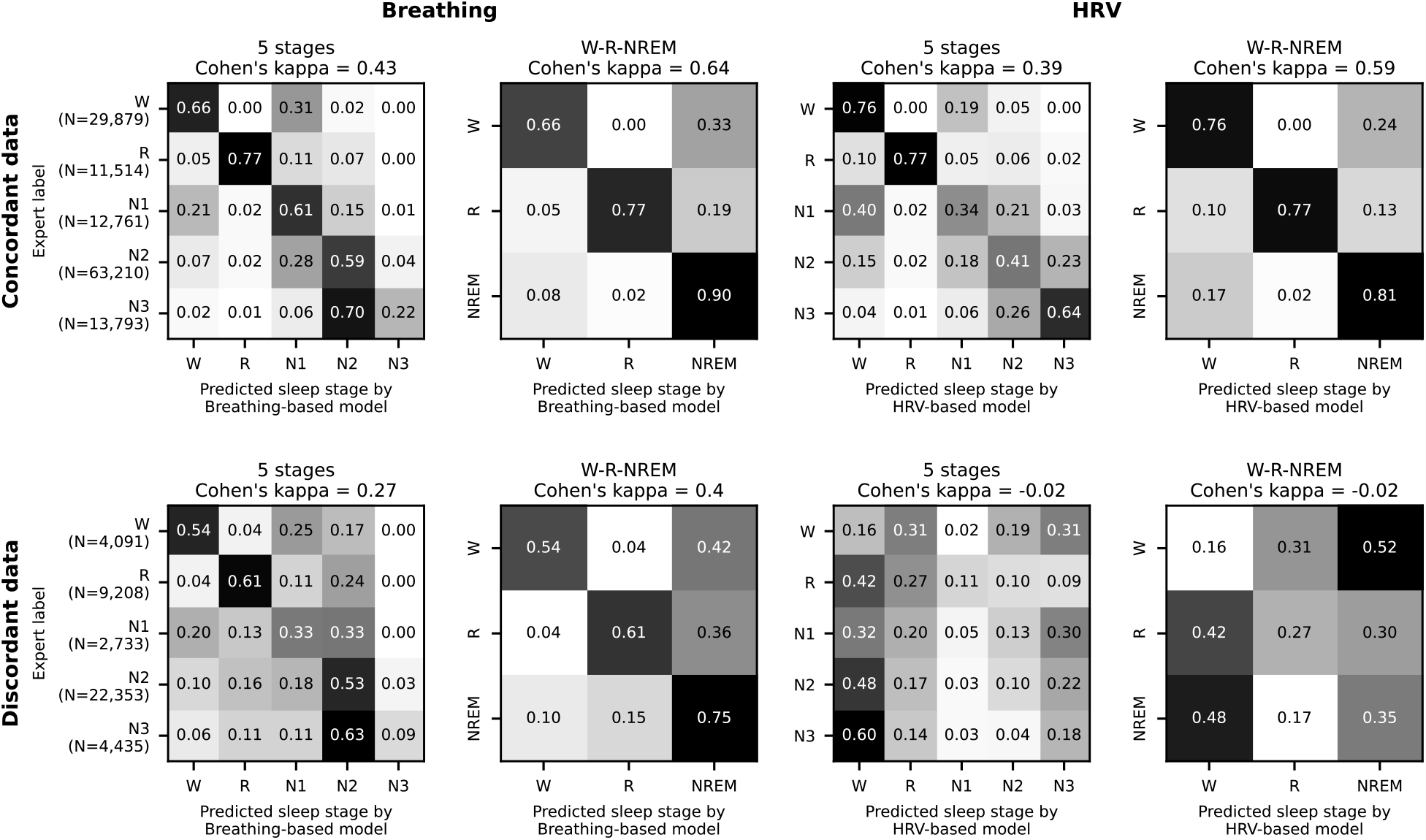
Model performance evaluation on 220 covariate-matched sleep laboratory patients. Data where the HRV- and breathing-based sleep stages were in concordance (see main text for definition) also showed higher agreement with human expert labels. In total, 75.4% of all 30-second epochs were assigned concordant sleep stages by the models. For the discordant sleep epochs, the breathing-based sleep stages had markedly higher agreement with the expert labels than the HRV-based sleep stages.

The three analysis approaches resulted in the following total sleep times (TST), concordant sleep times (CST), and proportions sleep (S(%)) per 24-hour segment (numbers given as mean (SD)):

**A1. Inclusion: <u>any sleep;</u> sleep indices computed on <u>total</u> sleep:** ICU: 102 subjects (274 24-hour segments), TST **6.2 (3.1)** hours, S(%) **50.4 (19.7)** Sleep lab: 220 subjects (220 segments), TST **4.9 (1.6)** hours, S(%) **73.9 (19.3)**
**A2. Inclusion: ≥<u>2 hours of concordant sleep</u>; sleep indices computed on <u>total</u> sleep**: ICU: 80 subjects (163 segments), TST **8.6 (3.0)** hours, S(%) **56.4 (18.7)** Sleep lab: 190 subjects (190 segments), TST **5.3 (1.3)** hours, S(%) **76.7 (16.4)**
**A3. Inclusion: <u>≥2 hours of concordant sleep</u>; sleep indices computed on <u>concordant</u> sleep:** ICU: 80 subjects (163 segments), CST **4.2 (1.7)** hours, S(%) **58.3 (26.9)** Sleep lab: 190 subjects (190 segments), CST **4.1 (1.4)** hours, S(%) **76.8 (18.2)**

Sleep staging results for each patient over time are shown in **Figure 2** and **Figure S2**, sample hypnograms for both ICU and sleep lab patients are shown in **Figure 3**.

**Figure 2.**
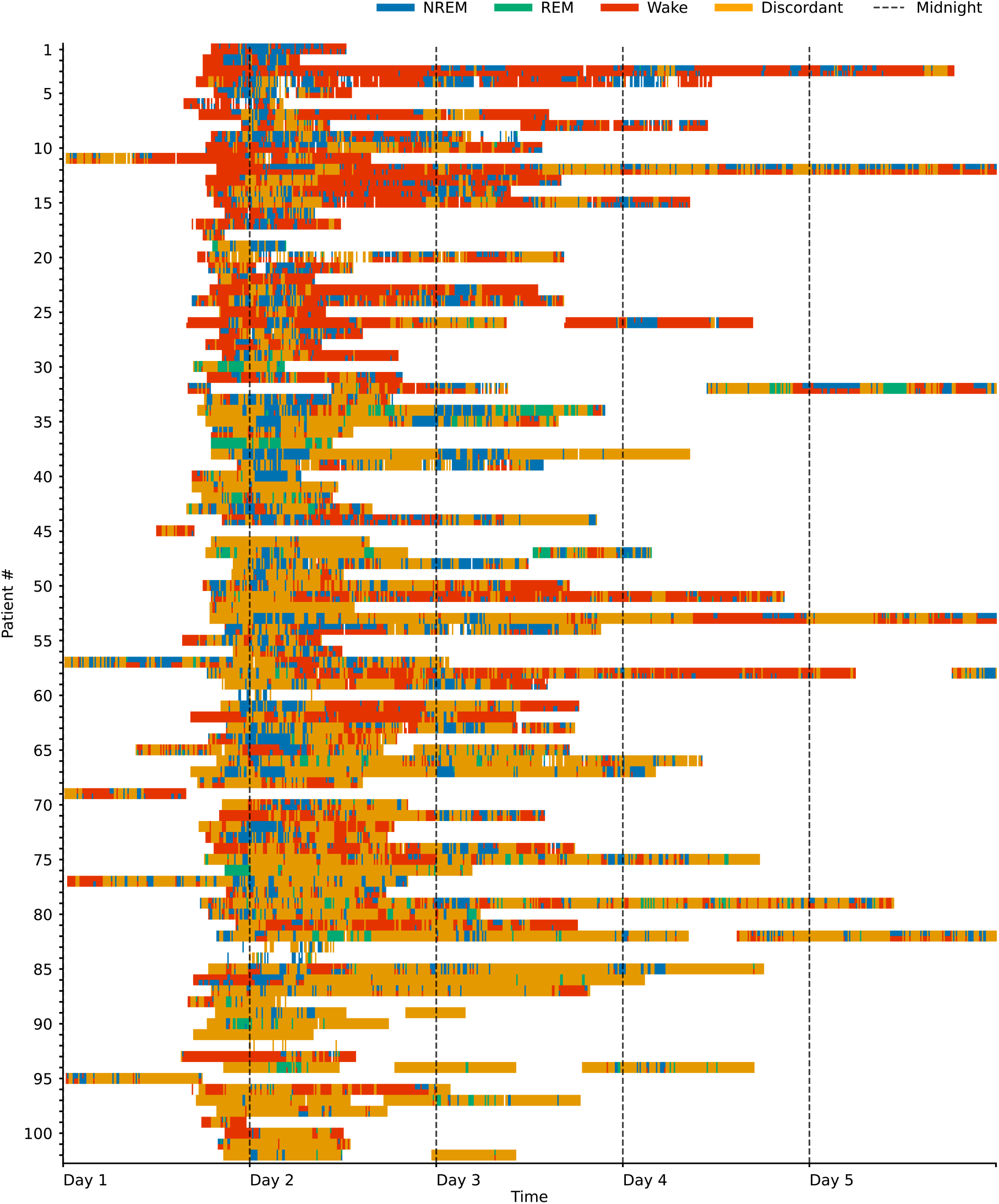
Swimmer plot visualizing sleep stages over time for 102 ICU patients. One line represents one patient, and patients are sorted by the proportion of sleep stage discordance (see main text for definition). For sleep epochs where HRV and breathing-based models were in concordance (60% of the data), the data is colored according to the sleep stages NREM (pooled N1, N2, N3), REM and Wake as assigned by the breathing-based (top half of each line) and HRV-based (bottom half of each line) sleep staging models. Epochs where HRV and breathing-based models were discordant (40% of the data) are marked as orange. Both sleep stage distribution and the amount of discordance considerably varied among patients.

**Figure 3.**
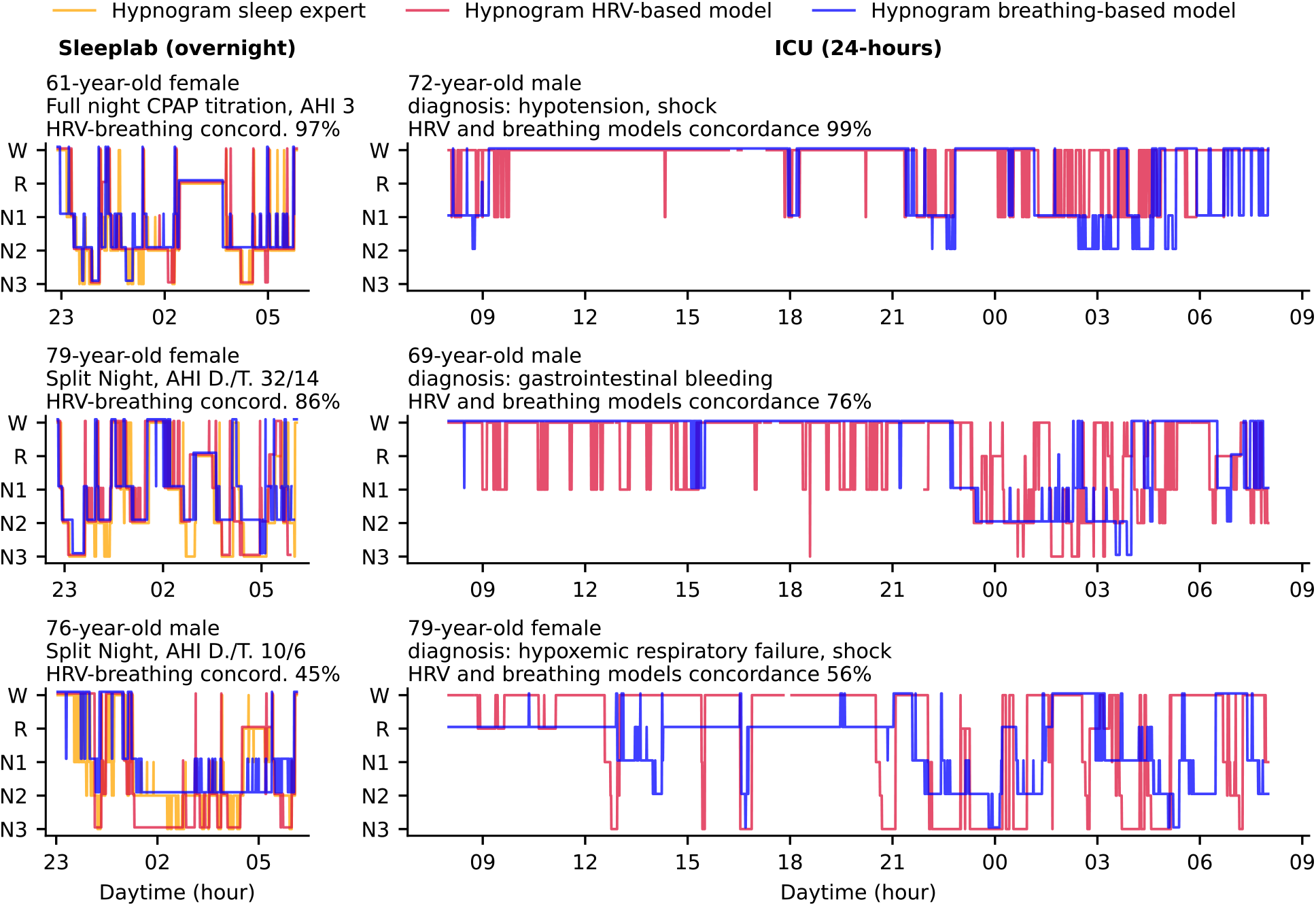
In this study, we used deep neural networks to stage sleep from both HRV- and breathing data, and defined concordance between the two resulting hypnograms (see main text). Hypnogram data for three sample patients from the sleep laboratory and three sample patients from the ICU are shown here and sorted by HRV- and breathing model concordance. The sleep laboratory data (clinical polysomnographies) included expert-scored sleep stages and respiratory events. Abbreviations: AHI D./T. Apnea-Hypopnea Index diagnostic/titration part of split night.

*Sleep indices.* For approach A2, the results of the sleep indices computation on a 24-hour level for the HRV and breathing models, together with the expert labels for the sleep lab data, are shown in **Figure 4**. Mean total sleep time per 24 hours in the ICU was determined to be 6.5 hours with the HRV-based model and 11.8 hours with the breathing-based model (+ 81.5%). In the sleep lab, total sleep time was determined to be 4.9 hours with HRV-based model and 5.7 hours with breathing model (+16.3%) and 5.6 hours by the human sleep expert.

**Figure 4.**
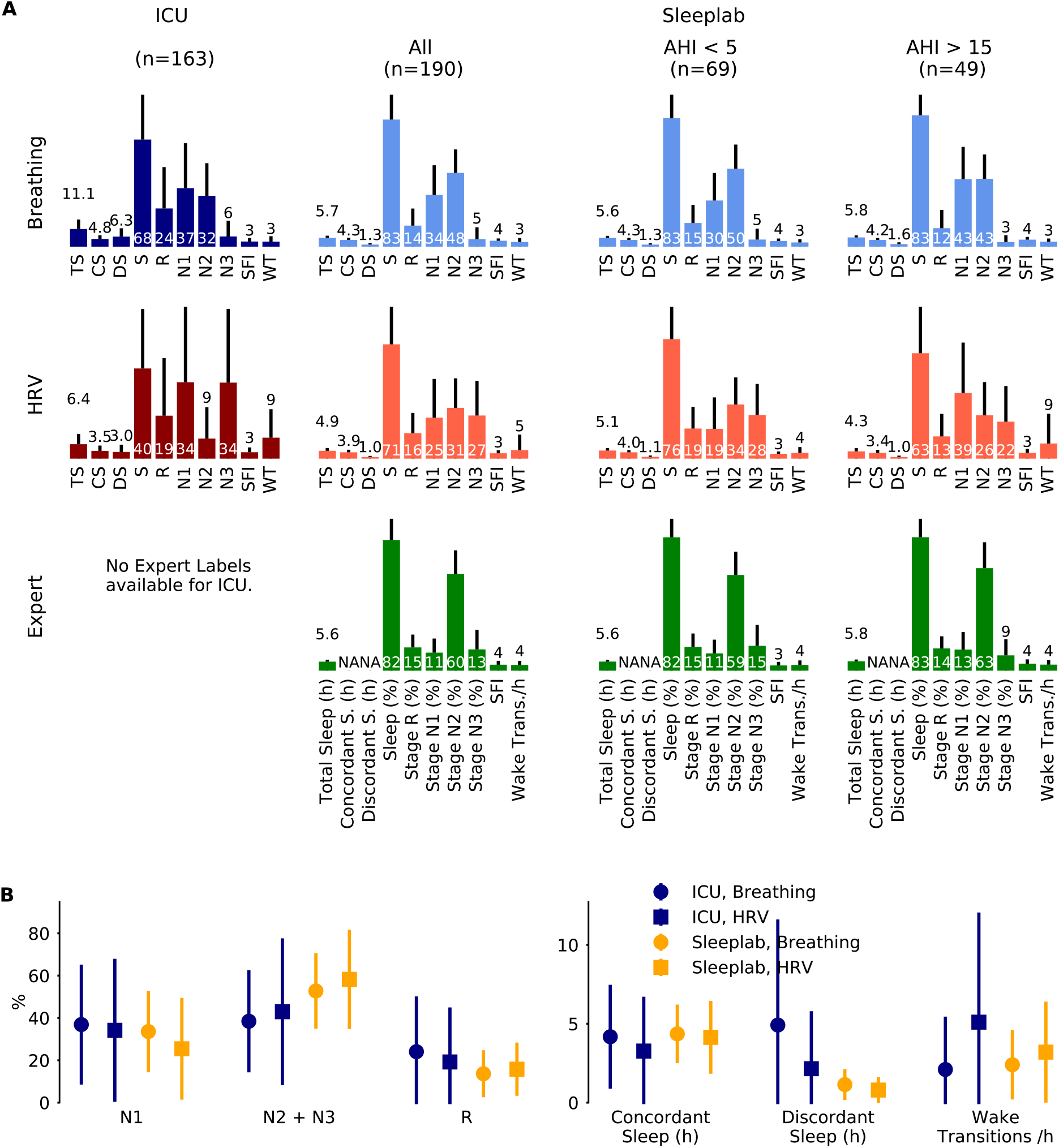
Sleep staging results for a surgical and medical ICU (N=80 subjects, 163 24-hour segments) and for an age- and sex-matched sleep laboratory cohort (N=190 subjects, 190 nights), where for each 24-hour segment or night sleep was detected at least once. Inclusion of all available ICU patients (N=102), i.e. without minimum sleep requirement, resulted in a mean total sleep time of 6.2 (3.1) hours per 24 hours in the ICU. Sleep stages were determined by breathing (respiratory effort) and heart rate variability (HRV)-based deep neural network models. For the sleep lab, additional human expert labels were available. **A.** Mean (one standard deviation) sleep indices. TS: total sleep time (hours), CS: concordant sleep time (hours), DS: discordant sleep time (hours), S: sleep percentage of total recording (%), SFI: sleep fragmentation index, WT: wake transitions per hour of sleep. **B.** Median (inter-quartile range) sleep indices for ICU and sleep lab cohort for both breathing- and HRV-based sleep staging models.

**Figure 5** depicts sleep indices (mean of breathing and HRV-based indices) distributions for all patients. In the ICU, there was a greater proportion of N1 compared to the AHI<5 sleep lab cohort (p MWU=0.05) and significantly less N2 compared to all sleep lab cohorts (p<0.001). The proportion of N2+N3 was reduced compared to total sleep lab (p<0.01) and sleep lab AHI<5 (p<0.001) cohorts but not compared to the sleep lab subgroup with AHI>15. While stage R distributions showed heavier tails in the ICU cohort (p MWU=0.05). Median wake transitions per hour of sleep were similar in the ICU and sleep lab AHI>15 groups (3.6 and 3.9), but lower in the sleep lab AHI<5 group (2.7, p<0.05). The ICU cohort showed a larger proportion of discordance between the breathing and HRV-based sleep staging models compared to the sleep lab cohort (41% and 19% respectively). In the ICU, a median of 25% of the day (08:00 – 20:00) and 41% of the night (20:00 – 08:00) were spent asleep. 38% of total sleep and 51% of R sleep occurred during daytime hours.

**Figure 5.**
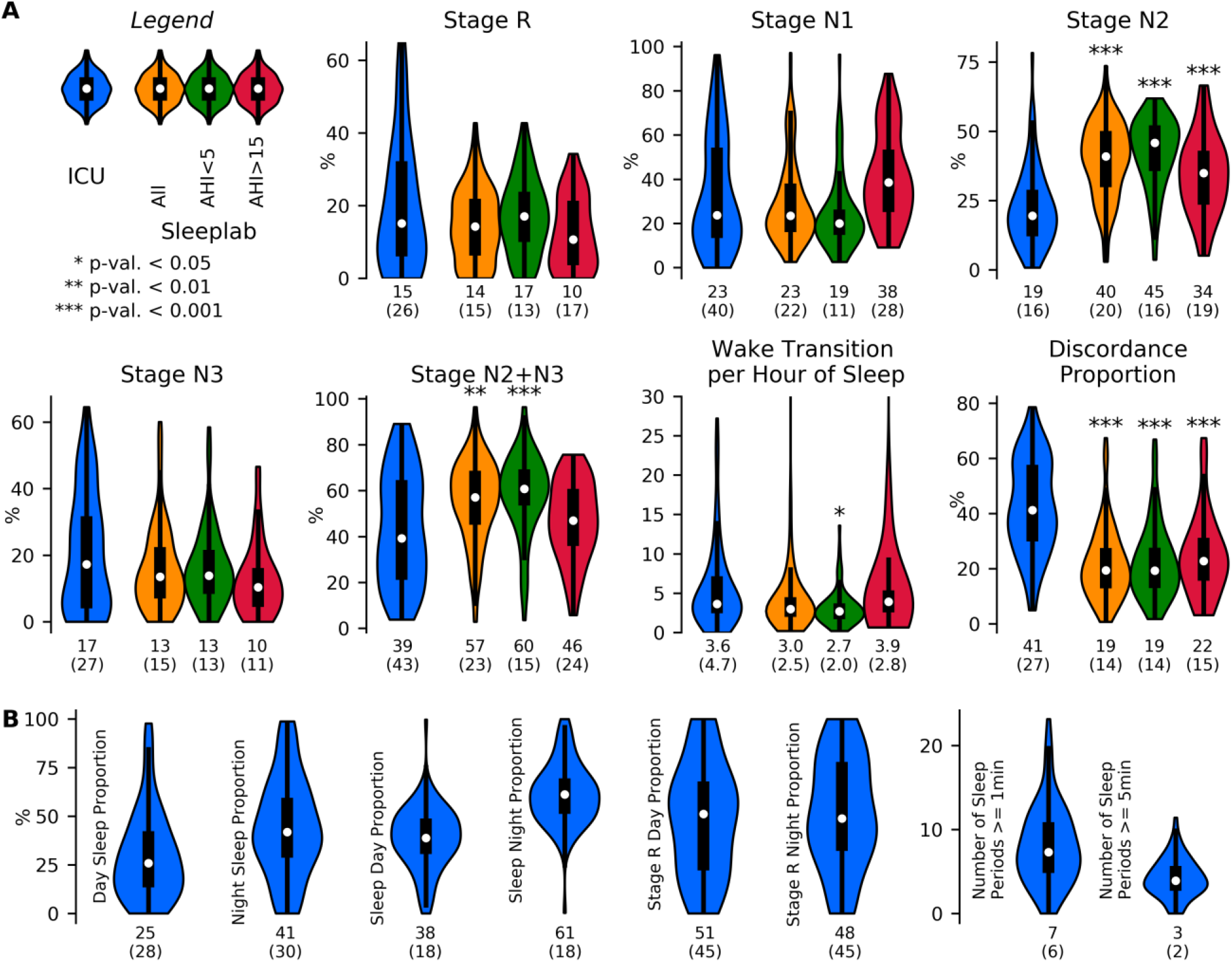
Sleep indices distribution visualized with violin plots and embedded boxplots (inter-quartile ranges: black rectangles) and medians (white dots); numerical values at bottom of distribution plots show median (interquartile range). **A.** Sleep Indices (mean of breathing and HRV-based indices) in the ICU (N=80 patients) and age and sex-matched sleep lab cohorts (N all = 190 subjects, N AHI<5 = 69 subjects, N AHI>15 = 49 subjects), both with requirement of minimum of 2 hours of detected sleep. Mann-Whitney U (MWU) tests and Mood’s median (MM) tests were applied to compare the ICU and sleep lab cohorts, and significance was indicated if both tests reached a given significance level. Stage R distributions showed heavier tails in the ICU cohort; statistical comparisons of differences did not reach significance. In the ICU, there was a larger proportion of light sleep (N1) compared to the AHI<5 sleep lab cohort (p MWU=0.05), and significantly less N2 compared to all sleep lab cohorts. The proportion of N2+N3 was also reduced compared to total sleep lab and AHI<5 cohorts but not compared to subjects with AHI>15. Median wake transitions per hour of sleep were similar in the ICU and AHI>15 group (3.6 and 3.9), and significantly lower in the AHI<5 group (2.7). The ICU cohort showed a significantly greater proportion of discordance between the breathing and HRV-based sleep staging models. **B.** Sleep fragmentation indices obtained in the ICU. Median 25% of the day (08:00 – 20:00) and 41% of the night (20:00 – 08:00) were spent asleep. 38% of total sleep and 51% of R sleep occurred during the day.

The following sleep indices results for the ICU were confirmed (same effect direction, all significant) by all analysis approaches A1-A3: Elevated discordant sleep time and proportion, reduced N2 proportion, and reduced N2+N3 proportion. The number of wake transitions per hour of sleep in the ICU was increased compared to the total sleep lab cohort (significant in 2 out of 3 analysis approaches), significantly increased compared to the sleep lab AHI<5 cohort (significant in all analysis approaches), and similar compared to the sleep lab AHI>15 cohort (not significantly different in any analysis approach). Results for the three analysis approaches are presented in **Tables S2-S6;** a summary is presented in **Table S7**.

*Subgroup Analysis.* The median and interquartile ranges of sleep indices for patients grouped by diagnosis are shown in **Figure 6** (and **Table S8** for numerical values including test results). Kruskal Wallis tests were not significant for any of the sleep indices (minimum p-value of 0.08 observed for N3 proportion). Patients with lowest N2+N3(%) observed had diagnoses of hemorrhage, shock, and encephalopathy, and patients with highest N1(%) had diagnoses of hemorrhage, sepsis and encephalopathy. Discordant sleep proportion was highest for patients with cirrhosis and liver transplant, pneumothorax, respiratory failure, and pneumonia, and lowest for patients with shock, hemorrhage, and acute kidney failure.

**Figure 6.**
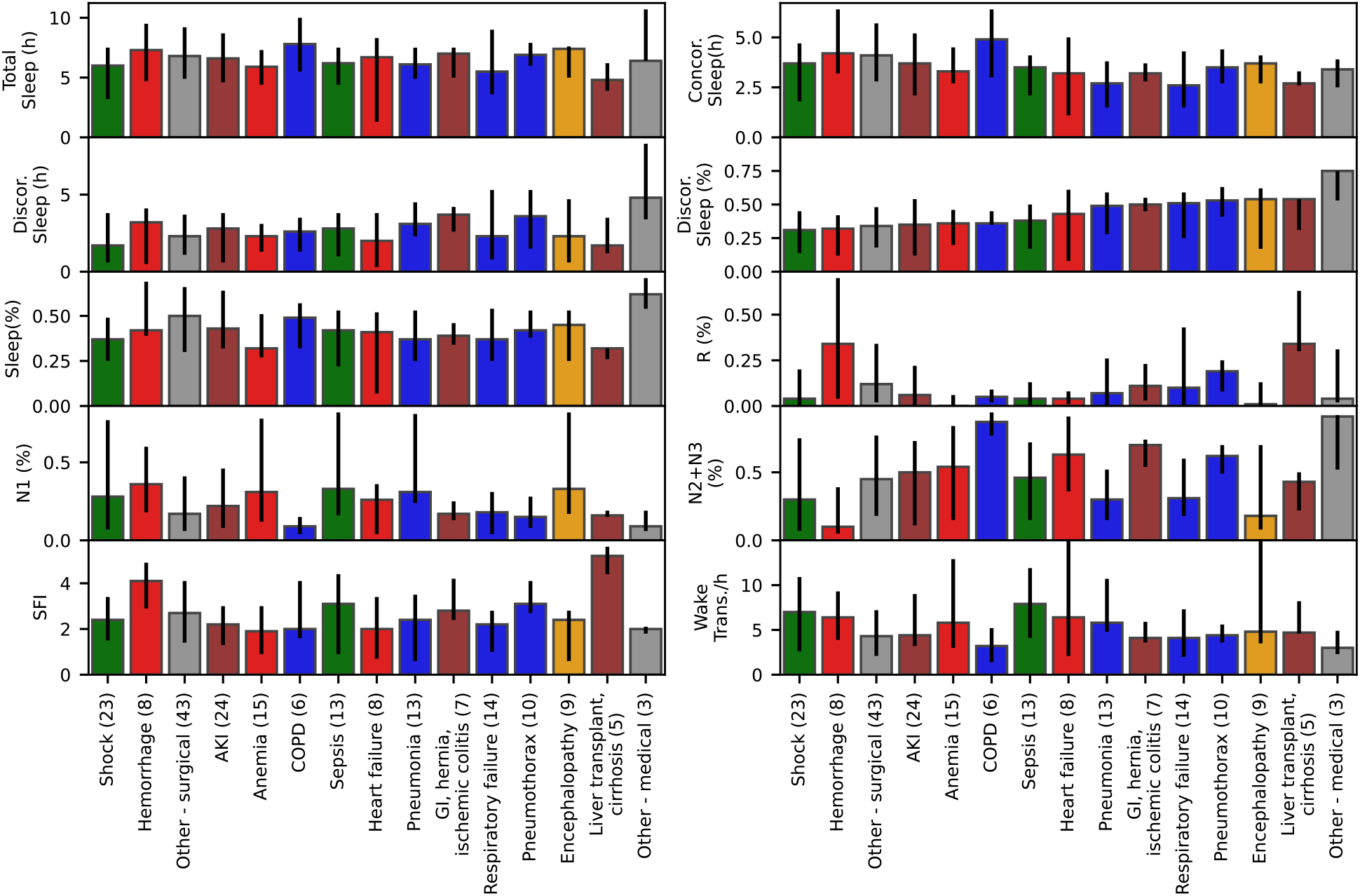
Median and interquartile ranges for sleep indices for ICU patients with at least two hours of detected sleep (N=80), grouped by their primary or main condition, with number of patients per condition shown in parentheses. Kruskal Wallis tests were not significant (p>0.05) for any of the sleep indices.

*Latent feature representation of sleep.* We generated 2-dimensional maps by using the deep neural network last hidden layer’s activation for each epoch as inputs for the UMAP (**Figure 7**). Coloring the data points with the predicted sleep stage shows clusters that correspond to those sleep stages (**Figure 7A**). For some sleep stages more than one cluster is apparent (HRV: R, N1, N3; Breathing: R, N3). For each sleep stage, estimated gaussian kernel densities of the UMAP-derived features show that the ICU features largely overlap with the sleep lab features, but are more widely dispersed (i.e., more variable) than in the sleep lab cohort (**Figure 7B**).

**Figure 7.**
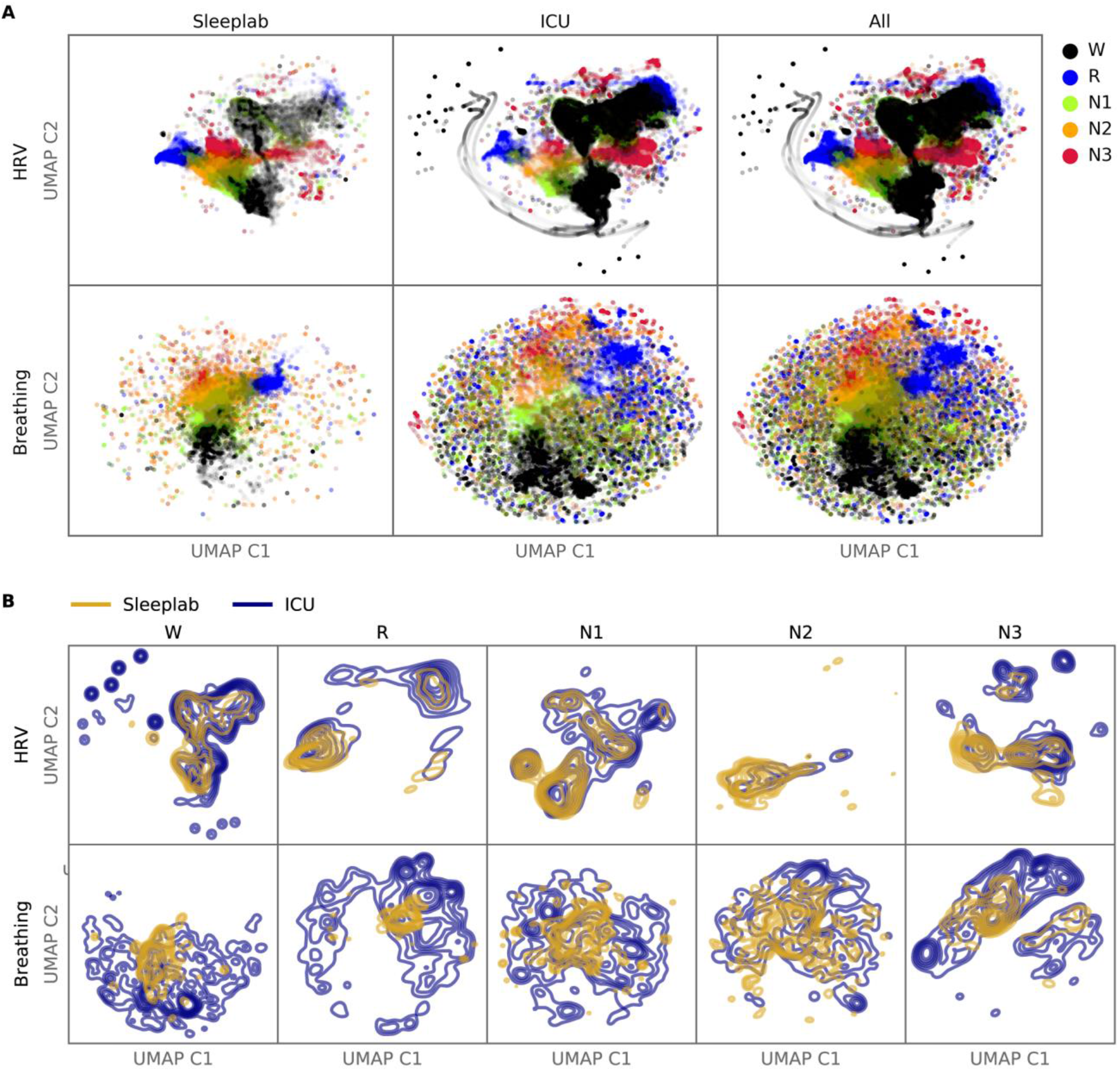
Latent Feature Analysis: Two-dimensional UMAP (Uniform Manifold Approximation and Projection) representations were computed with all epochs’ last hidden layer activations of the HRV-based and breathing-based deep neural networks. **A.** Data points are colored by predicted sleep stage, revealing clusters that correspond to sleep stages in both sleep lab and ICU data. **B.** For each sleep stage, probability density functions were estimated for the sleep lab and ICU data. While there is overlap between distributions (showing features computed from the ICU data are similar to features computed from the sleep lab data), the distributions of the ICU data are more dispersed (showing features in the ICU that are not present in the sleep lab data).

*Disagreement HRV and breathing model - error analysis.* Out of 44 computed biosignal-based features, 27 showed a significantly different distribution (Mann–Whitney U test, significance level 0.01) between concordant and discordant sleep, see **Tables S9-S10** for details on the difference between HRV and breathing models. For N1, significantly reduced mean values in discordant sleep were observed for HRV-VLF, HRV-LF, HRV-HF, HRV-RMSSD, inter-breath-intervals, respiratory variability, ventilation CVar, CPC-LFC, CPC-HFC, and significantly increased for respiratory rate. For N2, NN-interval duration, inter-breath-intervals, respiratory variability, ventilation CVar were reduced, while HRV-VLF, HRV-LF, and respiratory rate were increased. For N3, NN-interval duration was decreased, while HRV-VLF, HRV-LF, HRV-HF, HRV-RMSSD, respiratory variability, ventilation CVar and CPC-LFC were increased. Lastly, for REM sleep, we observed increased HRV-RMSSD and CPC-HFC for discordant sleep compared to concordant equivalents.

In the multivariate linear regression analysis, the automated feature selection (see supplement for details) led to a model with 53 input variables (F-test statistic of 1.51, a F-test p-value of 0.036) and an r-squared value of 0.424. Hence, on a 24-hour segment level, the computed HRV and breathing features could explain up to 42% of the variance of discordant sleep proportion.

None of SOFA scores, amounts of opioids, benzodiazepines or antipsychotics administered correlated with the discordant sleep proportion (all p-values for Pearson and Spearman correlations >0.05, see **Table S11**).

### G. Statistical Analysis – Breathing

**Figure 8** shows the mean and standard deviation of breathing features during sleep for each patient. The mean and standard deviation of respiratory rate during sleep was significantly larger for the ICU (mean respiratory rates = 17.4 cycles per minute, respiratory rate standard deviation = 4 cycles per minute, p<0.01), compared to the sleep lab patients. The median inter-breath-interval during sleep was significantly lower in the ICU (3.8 seconds) compared to the sleep lab (4.7 seconds). Effect directions were the same (with significance p<0.001) for the sleep lab AHI<5 and sleep lab AHI>15 groups. The mean and standard deviations of the ventilation’s coefficient of variation (as a proxy for minute ventilation) was significantly smaller in the ICU (p<0.05). The mean respiratory variability index in the ICU was smaller compared to the AHI>15 group (p<0.001) and larger to the AHI<5 group (not significant). See **Table S12** for a summary and **Table S13** for details.

**Figure 8.**
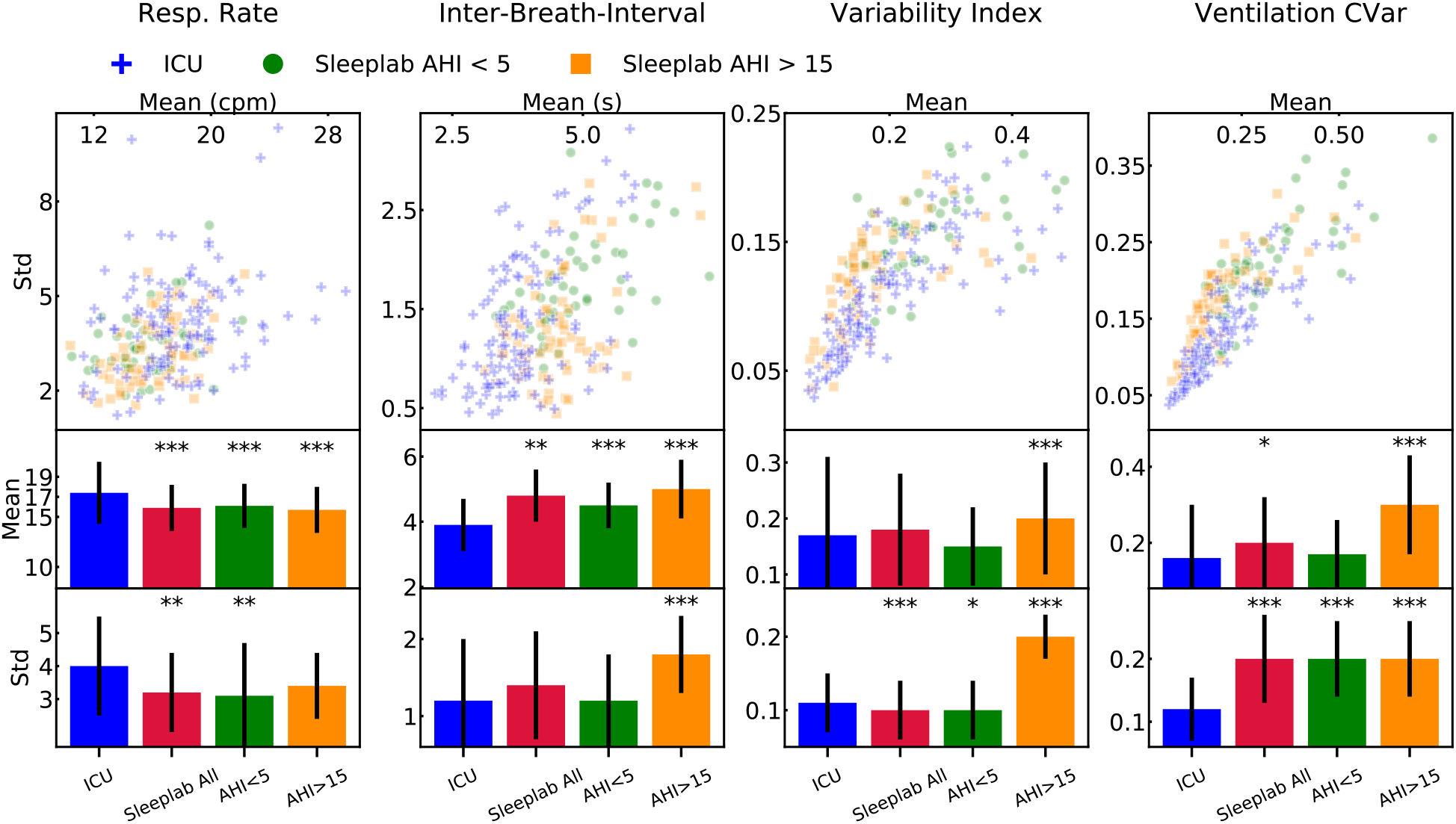
Breathing features during sleep for ICU (N=80) and sleep lab (N=190) patients. From the respiratory effort breathing signal obtained with a wearable band, we computed four features for all patients: respiratory rate, inter-breath-intervals, variability index, and as a proxy for minute ventilation, ventilation coefficient of variation (CVar). We compared the Intensive Care Unit (ICU) cohort with the full sleep lab cohort and the Apnea-Hypopnea-Index (AHI)-stratified cohorts. Significance levels are denoted as * for p<0.05 and *** for p<0.001. We show the scatter plots stratified by each sleep stage and tables containing the numerical values in the Online Supplement.

## IV. Discussion

Our main contributions are the following key findings: 1) In the ICU, heart rate variability (through ECG) and respiratory information provide meaningful information about sleep stage, quality, and fragmentation – this is significant because HRV and respiratory signals are much more easily obtained in the ICU than conventional polysomnography; 2) Clear differences are evident between sleep indices of ICU patients and those of an age and sex-matched clinical sleep laboratory cohort, such as decreased N2 and N2+N3 proportions in the ICU; 3) Sleep indices of ICU patients, particularly awakening frequency, resemble those of sleep-disordered breathing (SDB) sleep laboratory patients more closely than those of non-SDB patients; 4) Sleep stage concordance between HRV- and breathing-based models was associated with higher agreement with experts compared to sleep stage discordance; 5) The proportion of sleep stage concordance was lower in the ICU (60%) compared to the sleep laboratory cohort (81%); 6) Sleep in the ICU is fragmented; 7) ICU patients showed higher respiratory rate compared to sleep laboratory patients, higher breathing variability compared to non-SDB sleep laboratory patients, and lower breathing variability compared to SDB sleep laboratory patients.

*Sleep in the ICU* – In this study we compared sleep of ICU patients with covariate-matched sleep laboratory patients. Sleep indices of ICU patients obtained from the HRV- and breathing-based deep neural network sleep staging models were robust across sensitivity analyses. Compared to the sleep laboratory cohort, we observed a shift of NREM sleep stage proportion from N2 and N3 towards more light and non-restorative N1. This is significant, as there has been growing evidence that deep NREM sleep has a crucial role in neurophysiological phenomena such as immunity, glucose metabolism, hormone release and memory(49). The median amount of REM sleep in ICU patients was lower compared to non-sleep disordered breathing and similar to sleep disordered breathing patients from the sleep laboratory. Further, sleep of ICU patients was highly fragmented: patients spent a median of 25% of the day and 41% of the night asleep. Moreover, 38% of total sleep and 51% of REM sleep occurred during the day.

The significant differences between the ICU and the sleep laboratory patients provide strong evidence that sleep is frequently deranged during critical care. Our results reinforce prior published findings (typically PSG studies with smaller sample size(22)), including abnormal hypnograms, high arousal index, abnormal sleep stage shifts, a reduction of deep NREM and varying amount of REM sleep (22, 30, 50–53). Furthermore, there is growing evidence that sleep in the ICU, especially for ventilated patients, is often abnormal and cannot be scored with traditional EEG-based scoring criteria (18, 54). Potentially, periods of discordance observed in our study coincide with EEG patterns that have been classified as ‘atypical sleep’ and ‘pathological wakefulness’ in previous studies examining sleep in the ICU(18, 54).

The presented results are important as there is evidence that sleep impairment in the ICU is associated with delirium(19). Delirium, a state of brain dysfunction with fluctuating awareness, disorganized thinking, and an altered level of consciousness(19), is associated with long-lasting cognitive impairment after hospital discharge and accelerated onset of dementia(55). While the mechanism of delirium is not entirely established, the absence or reduction of deep, restorative NREM sleep and REM sleep, and disturbances of circadian rhythms may be risk factors of developing delirium (19). Cardiovascular and pulmonary signals, as we have shown here, can help monitoring sleep more routinely in the ICU, which can guide measures to preserve sleep continuity, such as managing noise and light levels better or concentrating times of procedures and exams to allow more consolidated periods of rest.

*Feasibility of staging sleep with heart rate variability and respiratory signals* – The deep neural network activation patterns, visualized in 2D-space with embedding maps, of ICU patients were mostly similar to those of sleep lab patients, with additional activation patterns only present in the ICU. This means that most of the ICU input data were processed by the artificial networks in such a way that the learned data representations (i.e., high-dimensional features about sleep before making the final sleep stage decision) resembled representations resulting from input data that are like the training data (the networks were trained using sleep lab data). This result helps mitigate concerns about covariance shift. Because sleep appears to be fundamentally changed in the ICU, ICU activation patterns that are not present in sleep lab patterns are potentially of interest, and more research is needed to understand the significance and implications of these clusters, as they may affect reliability of models or indicate certain pathological states present in the ICU population and not in sleep laboratory populations. 27 out of 44 HRV, CPC, and breathing features significantly differed between concordant and discordant sleep epochs, indicating that these well-established and interpretable features might be used a-priori to assess the reliability in sleep stage assessment by HRV- and breathing-based sleep staging models. This result was confirmed with a multivariate linear regression model, using HRV and breathing features from 24-hour segments as independent variables, where 42% of the variance of discordance proportion was explained by these features. Surprisingly, we found no association between the discordance of the models and use of opioids, benzodiazepines and antipsychotics, or health status of the patient (SOFA score). Similarly, patients grouped by primary and secondary diagnosis, did not show statistically significant differences in model discordance. This is remarkable, as we hypothesized more severe medical conditions would lead to a higher discordance, but non-significance may be a result of low sample size per subgroup.

These results demonstrate that the presented method of measuring sleep in ICU patients, insofar as our cohort is representative, is feasible in most cases, although it does lead to more inconclusive sleep stage assessments (measured by discordance of the HRV- and breathing-based sleep staging models) than in the sleep laboratory.

*Respiratory analysis* – Mean respiratory rate during sleep was significantly higher in the ICU compared to all sleep lab cohorts. Variance of respiratory rate was significantly increased compared to total sleep lab cohort and AHI<5 subgroup but not compared to AHI>15 subgroup. Conversely, mean inter-breath-intervals were significantly lower in the ICU compared to all sleep lab cohorts. Interestingly, the variance of the inter-breath-intervals was also low in the ICU (comparable to sleep lab AHI<5, significantly lower than AHI>15 group). Breathing variability, which considers variability in timing and amplitude of breaths, was higher in the ICU group than in the AHI<5 sleep lab patients and lower than in AHI>15 sleep lab patients. This suggests that there is greater baseline instability of respiratory patterns in the ICU, likely reflecting acute illness, even when respiration is mildly abnormal. On the other hand, this level of instability is seen in healthier (non-ICU) patients with severe sleep apnea. Similarities of breathing statistics in ICU patients and in SDB sleep laboratory patients, and overall higher breathing variability in the ICU patients, underscore the high prevalence of sleep-disordered breathing in the ICU patients.

*Limitations* – In our observational study, we did not have EEG recordings available in the ICU. Therefore, we could not analyze relationships of sleep EEG and the HRV- and breathing-based sleep stage assessments. Joint analysis of cerebral cortex, cardiac, and respiratory activity would allow a more holistic analysis and understanding of sleep physiology in the ICU but are a practical challenge in this population. While the sample size of our study is relatively high compared with other sleep studies done in the ICU setting, 102 patients is still likely a relatively low number relative to the full spectrum of ICU patients and sleep physiology. As the target enrollment of the clinical trial is 750 patients, we hope to be able to follow up with more granular analysis of sleep in the ICU in the future. Patients in this cohort were admitted to surgical and medical ICUs, thus results in other types of ICU’s (e.g., neurological, cardiothoracic) might be different. Further, because we studied sleep in patients mostly not mechanically ventilated, the accuracy of the proposed sleep measurement method and reported sleep results during mechanical ventilation might differ. Finally, all patients were randomized into three groups as part of our clinical trial, where they were infused overnight for 11 hours with 0.1 or 0.3 mcg/kg/h dexmedetomidine (low dose) or placebo (normal saline). When this ongoing quadruple blinded trial is concluded, it will be possible to analyze the effects of low dose dexmedetomidine, if any, on our measures of interest.

*Summary* – Studying sleep in the ICU is possible and repeatable with readily accessible biological signals: ECG and respiration, with an accuracy tradeoff compared to full polysomnography. However, the patterns noted may themselves encode information about disease pathologies in the ICU setting. HRV/respiration analysis may provide important physiological information not captured by EEG analysis. We found partial explanations for discordance in sleep stages determined by the HRV- and breathing-based models. Sleep architecture and breathing during sleep in the ICU differed from an age and sex-matched sleep lab cohort, including subgroups with and without disordered breathing. As expected, signatures of deep NREM sleep were reduced in the ICU, and dispersion of sleep into the day was noted. The ability to monitor sleep or sleep-like states in the ICU without complex monitoring can enable better tracking of sleep-wake state boundaries and fragmentation, the impact of such fragmentation on delirium and other ICU outcomes, and even estimate the effects of sleep targeted therapies.

## Data Availability

Data and models used in this research are available under: https://github.com/mghcdac/ecg_respiration_sleep_staging_icu/

## Author contribution

WG: data management, developed code and models, data analysis, drafted and reviewed the manuscript.

PVK: data acquisition, clinical chart review, classification of patients into diagnoses groups, drafted and reviewed the manuscript.

SQ: data acquisition, data management, critical review and revision of the manuscript.

RAT: data management, data acquisition, critical review and revision of the manuscript

AB: data management, data acquisition, critical review and revision of the manuscript

MDSC: was part of the data acquisition, critical review and revision of the manuscript.

NA: was part of data acquisition, critical review and revision of the manuscript.

MJL: was part of the data management, critical review and revision of the manuscript.

EMY: was part of the data management, critical review and revision of the manuscript.

AH: was part of data acquisition, critical review and revision of the manuscript.

SR: was part of data acquisition, critical review and revision of the manuscript.

EP: was part of data acquisition, critical review and revision of the manuscript.

LP: was part of data acquisition, critical review and revision of the manuscript.

JH: was part of data acquisition, critical review and revision of the manuscript.

MA: was part of data acquisition, critical review and revision of the manuscript.

BC: was part of the data acquisition, critical review and revision of the manuscript.

TH: critical review and revision of the manuscript

HS: critical review and revision of the manuscript.

YS: data management, critical review and revision of the manuscript.

SC: critical review and revision of the manuscript.

BTT: critical review and revision of the manuscript.

OA: critical review and revision of the manuscript.

DK: critical review and revision of the manuscript

RJT: design of the work, supervision, critical review and revision of the manuscript

MBW: design of the work, supervision, critical review and revision of the manuscript.

## Financial disclosure

Dr. Westover is a co-founder of Beacon Biosignals and reports grants from NIH, during the conduct of the study; Dr. Cash reports other COI from Neuralink, Paradromics, and Synchron, and Beacon Biosignals, outside the submitted work. Dr. Thomas reports personal fees from GLG Councils, Guidepoint Global, and Jazz Pharmaceutics, outside the submitted work. In addition, Dr. Thomas has a patent ECG-spectrogram with royalties paid by MyCardio, LLC, a patent Auto-CPAP with royalties paid by DeVilbiss-Drive, and an unlicensed patent CO2 device for central / complex sleep apnea issued. Dr. Thompson reports consulting for Bayer, Novartis, and Thetis, outside the submitted work. In addition, Mr. Kuller has a patent: Patent US10123724B2 “Breath volume monitoring system and method” issued.

## Non-financial disclosure

Mr. Kuller reports non-financial support from Myair Inc., during the conduct of the study, and non-financial support from Myair Inc, outside the submitted work.

Sources of Support (Grants/Gifts/Equipment and/or Drugs):

M.B.X was supported by the Glenn Foundation for Medical Research and American Federation for Aging Research (Breakthroughs in Gerontology Grant); American Academy of Sleep Medicine (AASM Foundation Strategic Research Award); Football Players Health Study (FPHS) at Harvard University; Department of Defense through a subcontract from Moberg ICU Solutions, Inc; and NIH (1R01NS102190, 1R01NS102574, 1R01NS107291, 1RF1AG064312).

## List of abbreviations

ECG: electrocardiogram
HRV: heart rate variability
ICU: intensive care unit
SOFA: sequential organ failure assessment
CCI: Charlson Comorbidity Index
TS: total sleep
CS: concordant sleep
DS: discordant sleep
S: sleep percentage
R: stage R
N1: stage N1
N2: stageN2
N3: stage N3
SFI: sleep fragmentation index
WT: wake transitions per hour of sleep
AHI: apnea hypopnea index
UMAP: Uniform Manifold Approximation and Projection
SDB: sleep disordered breathing
Std: standard deviation
IQR: inter quartile range
CVar: Coefficient of Variation
p_dag: p-value D’Agostino’s K-squared test
p_sha: p-value Shapiro Wilk test
p_tt: p-value Student’s t-test
s_tt: t-statistic Student’s t-test
p_mwu: p-value Mann Whitney U test
s_mwu: Mann Whitney U statistic
p_medians: p-value Mood’s median test
s_medians: Mood’s Median test statistic

## SUPPLEMENT

*A. Dataset –* Patients’ eligibility for the clinical trial was determined according to the following inclusion/exclusion criteria taken from the Investigation of Sleep in the Intensive Care Unit(1) ClinicalTrials.gov page:

Inclusion Criteria:

In order to be eligible to participate in this study, an individual must meet all of the following criteria:

1. Admitted to MGH Blake 7 or 12, or Ellison 4 ICU at Massachusetts General Hospital
2. Male or female, aged > 50 years.
3. Provision of signed and dated informed consent form (by patient or LAR).
4. Stated willingness to comply with all study procedures and availability for the duration of the study.
5. Not on mechanical ventilation at the time of enrollment.
6. Able to be enrolled before 7PM.
7. For females of reproductive potential: pregnancy test is negative.

Exclusion Criteria:

Any individual who meets any of the following criteria will be excluded from participation in this study:

1. Unable to be assessed for delirium (e.g. blindness or deafness).
2. Pregnancy or lactation.
3. Known allergic reactions to components of dexmedetomidine.
4. Follow-up would be difficult (e.g. active substance abuse, homelessness).
5. Severe dementia, as measured by a score of ≥3.3 on the Short Informant Questionnaire on Cognitive Decline in the Elderly (IQCODE).
6. Known pre-existing neurologic disease or injury with focal neurologic or cognitive deficits.
7. Serious cardiac disease (e.g. sick sinus syndrome, sinus bradycardia).
8. Severe liver dysfunction (Child-Pugh Class C).
9. Severe renal dysfunction (receiving dialysis).
10. Low likelihood of survival >24 hours.
11. Low likelihood of staying in the ICU overnight
12. Patient is receiving either of the anticholinergic drugs scopolamine or penehyclidine.
13. Concomitant enrollment in another study protocol that may interfere with data acquisition or reliability of measurements.
14. Deemed unsuitable for selection by the research team or ICU providers due to any medical, legal, social, or interpersonal issues that would either compromise the study or the routine care of patients.

**Table S1.** ICU and matched sleep laboratory cohort.

| Cohort | ICU | Sleeplab<br>All | Sleeplab<br>AHI < 5 | Sleeplab<br>AHI > 15 |
| --- | --- | --- | --- | --- |
| N Females | 41 | 98 | 40 | 18 |
| N Males | 62 | 122 | 37 | 34 |
| Ratio<br>Males/Females | 1.51 | 1.24 | 0.93 | 1.89 |
| Median Age<br>[IQR] | 68<br>[62.5,<br>75.0] | 67.3<br>[60.6,<br>73.4] | 66.0<br>[59.9, 69.5] | 70.5<br>[60.5,<br>74.8] |
| t-test Welch<br>p-value | - | 0.78 | 0.14 | 0.51 |
| Mann-Whitney U test<br>p-value | - | 0.78 | 0.15 | 0.49 |
| Median Age<br>Female [IQR] | 67<br>[57, 76] | 66.2<br>[59.9,<br>71.9] | 65.5<br>[58.8, 69.8] | 71.8<br>[67.3,<br>76.1] |
| t-test Welch p-value | - | 0.88 | 0.36 | 0.19 |
| Mann-Whitney U test<br>p-value | - | 0.87 | 0.36 | 0.15 |
| Median Age<br>Male [IQR] | 68<br>[64, 75] | 68.3<br>[61.8,<br>74.2] | 66.9<br>[62.1, 69.5] | 69.7<br>[60.1,<br>74.0] |
| t-test Welch p-value | - | 0.92 | 0.41 | 0.56 |
| Mann-Whitney U test<br>p-value | - | 0.92 | 0.40 | 0.56 |

**Figure S1.**
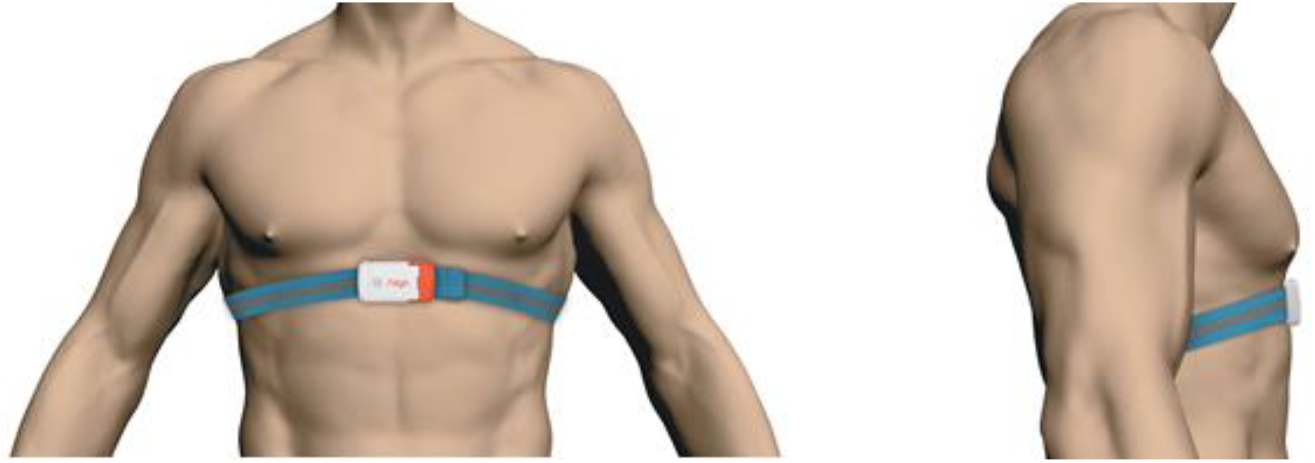
The wearable respiration device (‘Airgo’, MyAir LLC, Boston) used in this study.

### A. Biosignals Preprocessing

For ECG analysis, we mainly used the open source ‘PhysioNet Cardiovascular Signal Toolbox’ (2). The settings below correspond to the toolbox parameters we used for this study.

Window Size: 300 seconds windows; increment 30 seconds; minimum of 20% of data needs to be good quality for a window to be considered for analysis; maximum 15% of data is allowed to be missing in a window in order to be considered for analysis.

Signal Quality: low quality threshold = 0.9, comparison window length = 10 seconds; increment = 1 second; time threshold = 0.1 seconds; Margin time not to include in comparison = 2 seconds.

Preprocessing: Maximum believable gap in RR intervals = 2 seconds, Percent limit of change from one interval to the next = 100%; outlier method = ‘remove outlier points’, signal quality threshold for good data = 0.9; minimum length of good data = 30 seconds.

Frequency Domain Analysis Settings: ULF = [0 .0033]; VLF = [0.0033 .04]; LF = [.04 .15]; HF = [0.15 0.4]; Power Spectral Estimation Technique: Lomb Scargle.

Peak Detection Settings: Refectory period = 0.2 seconds; energy threshold = 0.15, window size for QRS detection = 15 seconds.

### B. Sleep Staging

**Figure S2.**
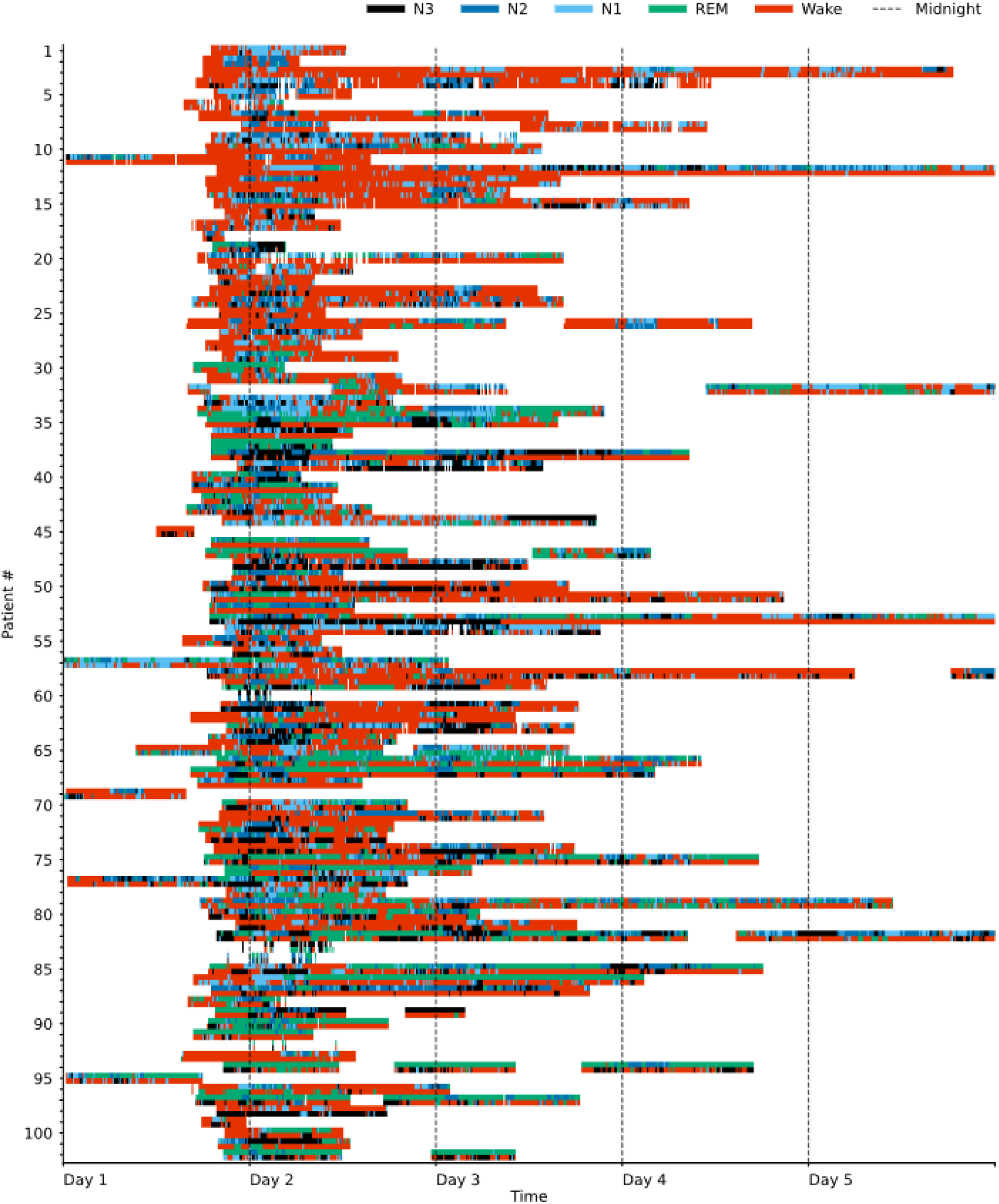
Swimmer plot visualizing sleep stages over time for 102 ICU patients. One line represents one patient and patients are sorted by the proportion of sleep stage discordance. The data is colored according to the sleep stages N1, N2, N3, REM and Wake as assigned by the breathing-based (top half of each line) and HRV-based (bottom half of each line) sleep staging models. Both sleep stage distribution and the amount of discordance considerably varied among patients.

**Figure S3.**
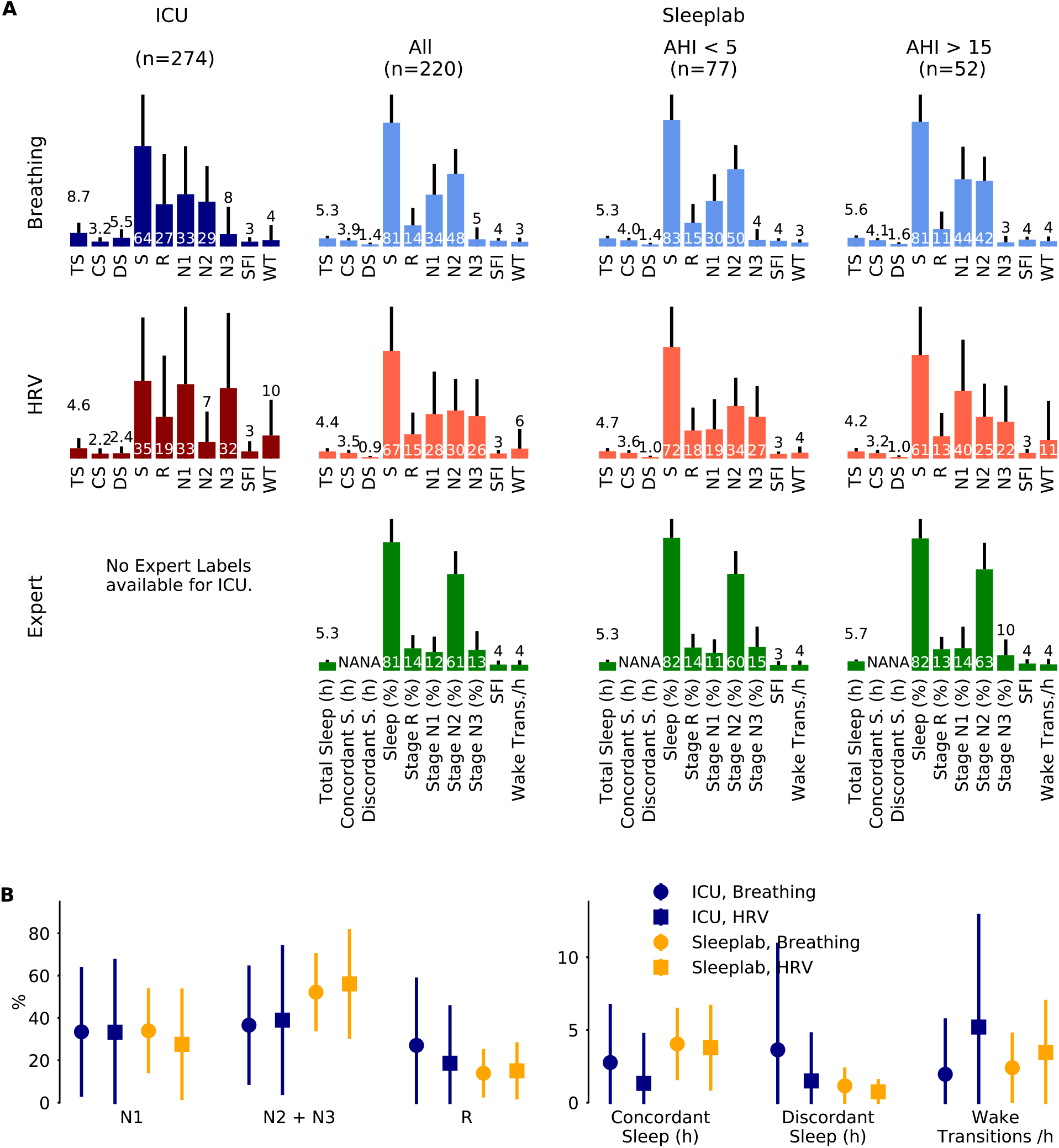
Analysis Approach A1. Sleep staging results for a surgical and medical ICU (N=102 subjects, 274 24-hour segments) and for an age and sex matched sleeplab cohort (N=220 subjects, 220 nights). Segments inclusion criteria: any sleep, sleep indices computed on total sleep. Sleep stages were determined by breathing (respiratory effort) and heart rate variability (HRV)-based deep neural network models, for the sleeplab additional human expert labels were available. **A.** Mean (one standard deviation) sleep indices for breathing, HRV and expert-based sleep stages (rows), and ICU, sleeplab and Apnea-Hypopnea Index (AHI) subgroups (columns). TS: total sleep time (hours), CS: concordant sleep time (hours), DS: discordant sleep time (hours), S: Sleep percentage of total recording (%), SFI: sleep fragmentation index, WT: wake transitions per hour of sleep. **B.** Median (inter-quartile range) sleep indices for ICU and sleeplab cohort for both breathing and HRV based sleep staging models.

**Figure S4.**
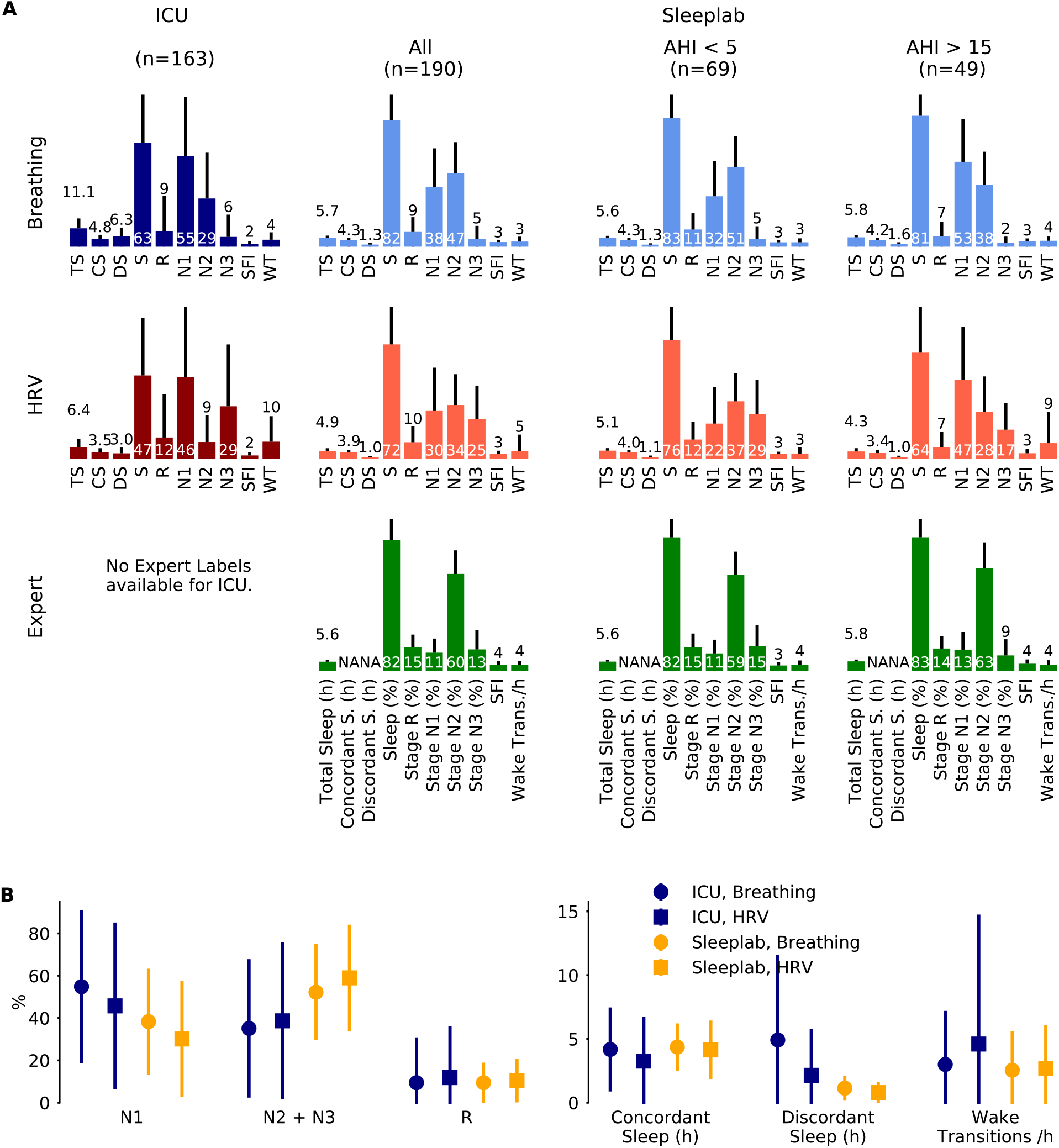
Analysis Approach A3. Sleep staging results for a surgical and medical ICU (N=80 subjects, 163 24-hour segments) and for an age and sex matched sleeplab cohort (N=190 subjects, 190 nights). Segments inclusion criteria: at least two hours of concordant sleep, sleep indices computed on concordant sleep. Sleep stages were determined by breathing (respiratory effort) and heart rate variability (HRV)-based deep neural network models, for the sleeplab additional human expert labels were available. **A.** Mean (one standard deviation) sleep indices for breathing, HRV and expert-based sleep stages (rows), and ICU, sleeplab and Apnea-Hypopnea Index (AHI) subgroups (columns). TS: total sleep time (hours), CS: concordant sleep time (hours), DS: discordant sleep time (hours), S: Sleep Percentage of total recording (%), SFI: sleep fragmentation index, WT: wake transitions per hour of sleep. **B.** Median (inter-quartile range) sleep indices for ICU and sleeplab cohort for both breathing and HRV based sleep staging models.

**Table S2.** Analysis approach A1. Summary of sleep stage analysis with average HRV and breathing-based sleep stage assessments. Inclusion all segments with any sleep, sleep statistics computed on total sleep.

|  | N | TS | CS | DS | DS (%) | S | R | N1 | N2 | N3 | N2+N3 | SFI | WT |
| --- | --- | --- | --- | --- | --- | --- | --- | --- | --- | --- | --- | --- | --- |
| ICU, Mean | 102 | 6.2 | 2.5 | 3.6 | 49.5 | 50.4 | 23.4 | 32.5 | 18 | 20 | 38 | 3.2 | 7 |
| ICU, Median | 102 | 6 | 2.3 | 3.4 | 48.2 | 51 | 17.4 | 29.2 | 17.5 | 15.2 | 35.2 | 2.8 | 5.3 |
| Lab All, Mean | 220 | ** 4.9 | ** 3.7 | ** 1.2 | ** 23.6 | ** 73.9 | ** 14.4 | 30.7 | ** 38.6 | ** 15.5 | ** 54.1 | 3.5 | ** 4.7 |
| Lab All, Median | 220 | 5.2 | ** 3.9 | ** 1.0 | ** 20.6 | ** 79.9 | 13.8 | 24.1 | ** 40.4 | 12.8 | ** 55.8 | 3.3 | ** 3.1 |
| Lab AHI 0-5, Mean | 77 | ** 5.0 | ** 3.8 | ** 1.2 | ** 23.7 | ** 77.1 | * 16.7 | ** 24.2 | ** 42.1 | 15.6 | ** 57.8 | 3.4 | ** 3.2 |
| Lab AHI 0-5, Median | 77 | 5.4 | ** 4.1 | ** 1.0 | ** 20.6 | ** 82.3 | 16.7 | ** 20.1 | ** 44.5 | 13.8 | ** 60.7 | 3.1 | ** 2.6 |
| Lab AHI 15-100, Mean | 52 | ** 4.9 | ** 3.7 | ** 1.3 | ** 25.3 | ** 71.0 | ** 12.3 | * 41.8 | ** 33.6 | ** 12.3 | * 45.9 | * 3.9 | 7.3 |
| Lab AHI 15-100, Median | 52 | 5.2 | ** 3.9 | ** 1.2 | ** 23.2 | ** 76.0 | 10.6 | 40.1 | ** 32.7 | 10 | * 46.1 | * 3.7 | 4 |
| ICU, Std |  | 3.1 | 1.7 | 2.4 | 20.4 | 19.7 | 20.6 | 22.1 | 12.7 | 17 | 20.3 | 2.1 | 6.7 |
| ICU, lqr |  | 5.2 | 2.3 | 3.4 | 30 | 31.8 | 32.3 | 31.1 | 14.8 | 25.4 | 30.8 | 2.1 | 5.4 |
| ICU, p_dag |  | 0.0322 | 0.0185 | 0.0363 | 0.4277 | 0.2723 | 0.0043 | 0.0216 | 0 | 0.0051 | 0.0521 | 0 | 0 |
| ICU, p_sha |  | 0.0525 | 0.0017 | 0.0005 | 0.5801 | 0.1666 | 0 | 0.0008 | 0 | 0 | 0.013 | 0 | 0 |
| ICU, gaussian |  | True | False | False | True | True | False | False | False | False | True | False | False |
| Lab All, p_tt |  | 0 | 0 | 0 | 0 | 0 | 0 | 0.483 | 0 | 0.0094 | 0 | 0.0738 | 0.0027 |
| Lab All, s_tt |  | 4.9 | -5.8 | 14 | 12.9 | -10.1 | 5.2 | 0.7 | -12.4 | 2.6 | -7 | -1.8 | 3 |
| Lab All, p_medians |  | 0.281 | 0 | 0 | 0 | 0 | 0.5492 | 0.0724 | 0 | 0.5492 | 0 | 0.1194 | 0.0007 |
| Lab All, s_medians |  | 1.2 | 29.1 | 57 | 76.5 | 72.3 | 0.4 | 3.2 | 98.9 | 0.4 | 34.5 | 2.4 | 11.5 |
| Lab All, p_mwu |  | 0.0005 | 0 | 0 | 0 | 0 | 0.0015 | 0.2619 | 0 | 0.0335 | 0 | 0.0012 | 0 |
| Lab All, s_mwu |  | 8647 | 6883.5 | 3670 | 3302 | 4391 | 8913 | 10724 | 3132 | 9796 | 6195 | 8857 | 7831.5 |
| Lab All, Std |  | 1.6 | 1.7 | 0.8 | 14.9 | 19.3 | 10.3 | 19.9 | 14.4 | 13.2 | 18.7 | 1.4 | 6.5 |
| Lab All, lqr |  | 2.3 | 2.6 | 0.9 | 17.1 | 27.9 | 15.6 | 22.8 | 20.7 | 16.4 | 25.2 | 1.7 | 3.1 |
| Lab All, p_dag |  | 0.0006 | 0 | 0 | 0 | 0 | 0.0169 | 0 | 0.0916 | 0 | 0.0066 | 0 | 0 |
| Lab All, p_sha |  | 0 | 0.0008 | 0 | 0 | 0 | 0 | 0 | 0.0246 | 0 | 0.0002 | 0.0001 | 0 |
| Lab All, gaussian |  | False | False | False | False | False | False | False | True | False | False | False | False |
| Lab AHI 0-5, p_tt |  | 0.0031 | 0 | 0 | 0 | 0 | 0.0101 | 0.0064 | 0 | 0.051 | 0 | 0.42 | 0 |
| Lab AHI 0-5, s_tt |  | 3 | -5 | 8.8 | 9.5 | -9.4 | 2.6 | 2.8 | -12.5 | 2 | -6.8 | -0.8 | 4.7 |
| Lab AHI 0-5, p_medians |  | 0.5899 | 0 | 0 | 0 | 0 | 0.9482 | 0.0011 | 0 | 0.5899 | 0 | 0.4244 | 0 |
| Lab AHI 0-5, s_medians |  | 0.3 | 21.1 | 51.6 | 51.6 | 41 | 0 | 10.6 | 77.8 | 0.3 | 41 | 0.6 | 17.3 |
| Lab AHI 0-5, p_mwu |  | 0.011 | 0 | 0 | 0 | 0 | 0.0961 | 0.0048 | 0 | 0.1485 | 0 | 0.0225 | 0 |
| Lab AHI 0-5, s_mwu |  | 3140 | 2220.5 | 1323 | 1141 | 1236 | 3479 | 3037 | 773 | 3568.5 | 1837 | 3238.5 | 2140 |
| Lab AHI 0-5, Std |  | 1.6 | 1.6 | 0.7 | 14.3 | 17.5 | 10.5 | 16.3 | 12.9 | 11.6 | 18 | 1.2 | 2.4 |
| Lab AHI 0-5, lqr |  | 2 | 2.2 | 0.9 | 15.1 | 23.1 | 15 | 12.5 | 16.5 | 12.9 | 17.6 | 1.5 | 2 |
| Lab AHI 0-5, p_dag |  | 0.0533 | 0.3946 | 0.0091 | 0.0002 | 0.0002 | 0.2759 | 0 | 0.0234 | 0 | 0.0053 | 0.4098 | 0 |
| Lab AHI 0-5, p_sha |  | 0.0086 | 0.2383 | 0.0001 | 0.0001 | 0 | 0.1244 | 0 | 0.0142 | 0 | 0.0013 | 0.1084 | 0 |
| Lab AHI 0-5, gaussian |  | True | True | False | False | False | True | False | False | False | False | True | False |
| Lab AHI 15-100, p_tt |  | 0.0073 | 0.0001 | 0 | 0 | 0 | 0.0003 | 0.0122 | 0 | 0.0032 | 0.0191 | 0.0243 | 0.85 |
| Lab AHI 15-100, s_tt |  | 2.7 | -3.9 | 7 | 7.6 | -6.3 | 3.7 | -2.5 | -6.9 | 3 | -2.4 | -2.3 | -0.2 |
| Lab AHI 15-100, p_medians |  | 0.3942 | 0.0012 | 0 | 0 | 0 | 0.1251 | 0.0609 | 0 | 0.0609 | 0.0106 | 0.0466 | 0.0609 |
| Lab AHI 15-100, s_medians |  | 0.7 | 10.5 | 35.6 | 35.6 | 18.1 | 2.4 | 3.5 | 27.9 | 3.5 | 6.5 | 4 | 3.5 |
| Lab AHI 15-100, p_mwu |  | 0.0198 | 0.0001 | 0 | 0 | 0 | 0.0014 | 0.0033 | 0 | 0.0062 | 0.005 | 0.0005 | 0.0872 |
| Lab AHI 15-100, s_mwu |  | 2113 | 1635 | 982 | 887 | 1136 | 1869 | 1940 | 1018 | 1997 | 1977 | 1795 | 2296 |
| Lab AHI 15-100, Std |  | 1.4 | 1.5 | 0.8 | 14.8 | 18.5 | 9.8 | 20.6 | 14.5 | 11 | 17.7 | 1.6 | 11.8 |
| Lab AHI 15-100, lqr |  | 1.6 | 2.1 | 0.8 | 15.5 | 25.1 | 17.1 | 37.6 | 19.5 | 11.9 | 24.3 | 1.8 | 2.9 |
| Lab AHI 15-100, p_dag |  | 0.1139 | 0.2976 | 0.0001 | 0.0114 | 0.0358 | 0.0299 | 0.075 | 0.8121 | 0.0004 | 0.2887 | 0.0122 | 0 |
| Lab AHI 15-100, p_sha |  | 0.034 | 0.2246 | 0.0011 | 0.0072 | 0.0033 | 0.0034 | 0.029 | 0.8231 | 0.0001 | 0.1343 | 0.015 | 0 |
| Lab AHI 15-100, gaussian |  | True | True | False | False | False | False | True | True | False | True | False | False |

**Table S3.** Analysis approach A2. Summary of sleep stage analysis with average HRV and breathing-based sleep stage assessments. Inclusion > 2 hours of concordant sleep, sleep statistics computed on total sleep.

|  | N | TS | CS | DS | DS (%) | S | R | N1 | N2 | N3 | N2+N3 | SFI | WT |
| --- | --- | --- | --- | --- | --- | --- | --- | --- | --- | --- | --- | --- | --- |
| ICU, Mean | 80 | 8.6 | 4.2 | 4.4 | 42.7 | 56.4 | 21.9 | 33.5 | 22.2 | 21 | 43.2 | 2.9 | 5.6 |
| ICU, Median | 80 | 8.2 | 4 | 3.9 | 41.2 | 58.2 | 15.1 | 23.7 | 19.5 | 17.2 | 39.3 | 2.9 | 3.6 |
| Lab All, Mean | 190 | ** 5.3 | 4.1 | ** 1.1 | ** 21.6 | ** 76.7 | ** 14.7 | 29.5 | ** 39.7 | ** 15.8 | ** 55.5 | ** 3.6 | * 4.2 |
| Lab All, Median | 190 | ** 5.5 | 4.2 | ** 1.0 | ** 19.4 | ** 81.1 | 14.2 | 23.5 | ** 40.9 | 13.5 | ** 57.1 | * 3.4 | 3 |
| Lab AHI 0-5, Mean | 69 | ** 5.4 | 4.2 | ** 1.2 | ** 22.4 | ** 79.4 | 17.2 | * 24.3 | ** 42.4 | 16.1 | ** 58.5 | 3.4 | ** 3.2 |
| Lab AHI 0-5, Median | 69 | ** 5.7 | 4.3 | ** 1.0 | ** 19.3 | ** 84.2 | 17.1 | 19.9 | ** 45.8 | 13.9 | ** 60.8 | 3.1 | * 2.7 |
| Lab AHI 15-100, Mean | 49 | ** 5.1 | 3.8 | ** 1.3 | ** 24.6 | ** 72.8 | ** 12.4 | 40.8 | ** 34.3 | ** 12.5 | 46.7 | ** 3.9 | 6.1 |
| Lab AHI 15-100, Median | 49 | ** 5.3 | 4 | ** 1.2 | ** 22.8 | ** 78.4 | 10.6 | 38.6 | ** 34.9 | 10.4 | 47 | 3.6 | 3.9 |
| ICU, Std |  | 3 | 1.7 | 2.5 | 16.9 | 18.7 | 21.3 | 25.5 | 14 | 17.9 | 24.1 | 1.4 | 5.4 |
| ICU, Iqr |  | 4.3 | 2.6 | 3.7 | 27.8 | 24.6 | 26.2 | 40.9 | 16.8 | 27.6 | 43.5 | 2 | 4.7 |
| ICU, p_dag |  | 0.0051 | 0.0603 | 0.1126 | 0.1981 | 0.5185 | 0 | 0.0162 | 0 | 0.0098 | 0 | 0.0002 | 0 |
| ICU, p_sha |  | 0.0261 | 0.0037 | 0.0184 | 0.2501 | 0.1902 | 0 | 0.0001 | 0.0001 | 0 | 0.006 | 0.0012 | 0 |
| ICU, gaussian |  | False | True | True | True | True | False | False | False | False | False | False | False |
| Lab All, p_tt |  | 0 | 0.5921 | 0 | 0 | 0 | 0.0002 | 0.1611 | 0 | 0.007 | 0 | 0.0008 | 0.0388 |
| Lab All, s_tt |  | 12.7 | 0.5 | 16.2 | 11.1 | -8.9 | 3.8 | 1.4 | -9.6 | 2.7 | -4.6 | -3.4 | 2.1 |
| Lab All, p_medians |  | 0 | 0.5052 | 0 | 0 | 0 | 1 | 1 | 0 | 0.3508 | 0.0051 | 0.0456 | 0.07 |
| Lab All, s_medians |  | 42.6 | 0.4 | 75 | 61.8 | 49.9 | 0 | 0 | 57.7 | 0.9 | 7.8 | 4 | 3.3 |
| Lab All, p_mwu |  | 0 | 0.4646 | 0 | 0 | 0 | 0.0535 | 0.3888 | 0 | 0.0716 | 0 | 0.0002 | 0.0064 |
| Lab All, s_mwu |  | 2558.5 | 7547.5 | 1440 | 2389 | 3071 | 6655 | 7434 | 2658 | 6742 | 5276 | 5547.5 | 6141.5 |
| Lab All, Std |  | 1.3 | 1.4 | 0.7 | 13 | 16.4 | 10.1 | 19 | 13.6 | 12.5 | 18 | 1.3 | 5.3 |
| Lab All, Iqr |  | 1.9 | 2.3 | 0.8 | 14.7 | 22.3 | 15.7 | 22 | 20.3 | 15.5 | 23.5 | 1.6 | 2.5 |
| Lab All, p_dag |  | 0.0258 | 0.0004 | 0 | 0 | 0 | 0.0358 | 0 | 0.0826 | 0 | 0.0036 | 0 | 0 |
| Lab All, p_sha |  | 0.0033 | 0.0149 | 0 | 0 | 0 | 0.0001 | 0 | 0.0212 | 0 | 0.0006 | 0.0002 | 0 |
| Lab All, gaussian |  | False | False | False | False | False | False | False | True | False | False | False | False |
| Lab AHI 0-5, p_tt |  | 0 | 0.805 | 0 | 0 | 0 | 0.0948 | 0.0122 | 0 | 0.0539 | 0 | 0.0555 | 0.0006 |
| Lab AHI 0-5, s_tt |  | 8.3 | 0.2 | 10.1 | 8.1 | -8.3 | 1.7 | 2.5 | -9.2 | 1.9 | -4.4 | -1.9 | 3.5 |
| Lab AHI 0-5, p_medians |  | 0 | 0.4634 | 0 | 0 | 0 | 0.6857 | 0.1171 | 0 | 0.363 | 0.0008 | 0.3758 | 0.0107 |
| Lab AHI 0-5, s_medians |  | 51.2 | 0.5 | 56 | 38 | 39.9 | 0.2 | 2.5 | 53.4 | 0.8 | 11.3 | 0.8 | 6.5 |
| Lab AHI 0-5, p_mwu |  | 0 | 0.3609 | 0 | 0 | 0 | 0.4297 | 0.0494 | 0 | 0.186 | 0.0001 | 0.0162 | 0.0006 |
| Lab AHI 0-5, s_mwu |  | 961 | 2666 | 557 | 925 | 914 | 2713 | 2326 | 786 | 2525 | 1744 | 2197.5 | 1904.5 |
| Lab AHI 0-5, Std |  | 1.2 | 1.3 | 0.7 | 13.4 | 14.7 | 10.4 | 16.9 | 12.7 | 11.5 | 17.6 | 1.2 | 2.4 |
| Lab AHI 0-5, Iqr |  | 1.8 | 1.9 | 0.7 | 14.7 | 16.8 | 13.9 | 11.6 | 16.5 | 13.2 | 15.8 | 1.5 | 2 |
| Lab AHI 0-5, p_dag |  | 0.435 | 0.8154 | 0.0085 | 0.0001 | 0.0079 | 0.3745 | 0 | 0.0052 | 0 | 0.0026 | 0.4507 | 0 |
| Lab AHI 0-5, p_sha |  | 0.2551 | 0.8811 | 0.0004 | 0.0002 | 0 | 0.2211 | 0 | 0.0015 | 0 | 0.0017 | 0.1325 | 0 |
| Lab AHI 0-5, gaussian |  | True | True | False | False | False | True | False | False | False | False | True | False |
| Lab AHI 15-100, p_tt |  | 0 | 0.1552 | 0 | 0 | 0 | 0.0041 | 0.0901 | 0 | 0.0031 | 0.3668 | 0.0005 | 0.7434 |
| Lab AHI 15-100, s_tt |  | 7.8 | 1.4 | 8.3 | 6.2 | -5 | 2.9 | -1.7 | -4.7 | 3 | -0.9 | -3.6 | -0.3 |
| Lab AHI 15-100, p_medians |  | 0 | 0.9451 | 0 | 0 | 0.0002 | 0.3079 | 0.0597 | 0 | 0.1669 | 0.2471 | 0.0597 | 0.8362 |
| Lab AHI 15-100, s_medians |  | 46.6 | 0 | 57 | 28.9 | 13.7 | 1 | 3.5 | 16.5 | 1.9 | 1.3 | 3.5 | 0 |
| Lab AHI 15-100, p_mwu |  | 0 | 0.1524 | 0 | 0 | 0 | 0.0165 | 0.0108 | 0 | 0.0155 | 0.1391 | 0.0003 | 0.4316 |
| Lab AHI 15-100, s_mwu |  | 624 | 1748 | 435 | 786 | 951 | 1520 | 1486 | 985 | 1515 | 1736 | 1248 | 1924 |
| Lab AHI 15-100, Std |  | 1.3 | 1.5 | 0.8 | 14.7 | 17.2 | 10 | 20.6 | 14.5 | 10.8 | 17.3 | 1.6 | 9.2 |
| Lab AHI 15-100, Iqr |  | 1.3 | 1.8 | 0.8 | 15.5 | 19.3 | 17.8 | 28.2 | 19.4 | 11.8 | 24.9 | 1.8 | 2.8 |
| Lab AHI 15-100, p_dag |  | 0.0595 | 0.4443 | 0 | 0.0048 | 0.0129 | 0.0308 | 0.136 | 0.8992 | 0.0002 | 0.3675 | 0.013 | 0 |
| Lab AHI 15-100, p_sha |  | 0.034 | 0.2717 | 0.0005 | 0.0042 | 0.004 | 0.0046 | 0.0306 | 0.8752 | 0.0001 | 0.1464 | 0.0139 | 0 |
| Lab AHI 15-100, gaussian |  | True | True | False | False | False | False | True | True | False | True | False | False |

**Table S4.** Analysis approach A3. Summary of sleep stage analysis with average HRV and breathing-based sleep stage assessments. Inclusion > 2 hours of concordant sleep, sleep statistics computed on concordant sleep.

|  | N | TS | CS | DS | DS (%) | S | R | N1 | N2 | N3 | N2+N3 | SFI | WT |
| --- | --- | --- | --- | --- | --- | --- | --- | --- | --- | --- | --- | --- | --- |
| ICU, Mean | 80 | 8.6 | 4.2 | 4.4 | 42.7 | 58.3 | 12.1 | 46.2 | 21.5 | 18.8 | 40.2 | 1.7 | 6.4 |
| ICU, Median | 80 | 8.2 | 4 | 3.9 | 41.2 | 54.8 | 0 | 37.6 | 19.3 | 11.1 | 33.6 | 1.3 | 4.1 |
| Lab All, Mean | 190 | ** 5.3 | 4.1 | ** 1.1 | ** 21.6 | ** 76.8 | 9.9 | ** 34.2 | ** 40.6 | 15 | ** 55.6 | ** 3.0 | ** 4.1 |
| Lab All, Median | 190 | ** 5.5 | 4.2 | ** 1.0 | ** 19.4 | ** 81.6 | ** 7.8 | ** 25.7 | ** 43.6 | 12.5 | ** 58.7 | ** 2.9 | * 2.6 |
| Lab AHI 0-5, Mean | 69 | ** 5.4 | 4.2 | ** 1.2 | ** 22.4 | ** 79.5 | 11.6 | ** 27.5 | ** 44.1 | 16.8 | ** 60.9 | ** 2.8 | ** 3.1 |
| Lab AHI 0-5, Median | 69 | ** 5.7 | 4.3 | ** 1.0 | ** 19.3 | ** 84.1 | ** 9.3 | ** 20.9 | ** 47.9 | 14.9 | ** 64.5 | ** 2.8 | ** 2.1 |
| Lab AHI 15-100, Mean | 49 | ** 5.1 | 3.8 | ** 1.3 | ** 24.6 | ** 72.6 | 6.7 | 50.1 | ** 33.2 | ** 9.9 | 43.1 | ** 3.3 | 6.5 |
| Lab AHI 15-100, Median | 49 | ** 5.3 | 4 | ** 1.2 | ** 22.8 | ** 75.5 | 1.6 | 47.5 | ** 35.1 | 7.6 | 46.1 | ** 3.0 | 4 |
| ICU, Std |  | 3 | 1.7 | 2.5 | 16.9 | 26.9 | 24.3 | 33.8 | 17 | 20.6 | 32.3 | 1.5 | 8.1 |
| ICU, Iqr |  | 4.3 | 2.6 | 3.7 | 27.8 | 42.4 | 11.9 | 58.1 | 25.7 | 31.3 | 63.5 | 2 | 6.6 |
| ICU, p_dag |  | 0.0051 | 0.0603 | 0.1126 | 0.1981 | 0.0001 | 0 | 0 | 0.0717 | 0.0071 | 0 | 0 | 0 |
| ICU, p_sha |  | 0.0261 | 0.0037 | 0.0184 | 0.2501 | 0.0026 | 0 | 0 | 0.0008 | 0 | 0 | 0 | 0 |
| ICU, gaussian |  | False | True | True | True | False | False | False | True | False | False | False | False |
| Lab All, p_tt |  | 0 | 0.5921 | 0 | 0 | 0 | 0.2904 | 0.0013 | 0 | 0.0731 | 0 | 0 | 0.0092 |
| Lab All, s_tt |  | 12.7 | 0.5 | 16.2 | 11.1 | -6.6 | 1.1 | 3.2 | -8.7 | 1.8 | -4.4 | -7 | 2.6 |
| Lab All, p_medians |  | 0 | 0.5052 | 0 | 0 | 0 | 0.0001 | 0.0051 | 0 | 0.6893 | 0.0022 | 0 | 0.0377 |
| Lab All, s_medians |  | 42.6 | 0.4 | 75 | 61.8 | 21.8 | 14.9 | 7.8 | 46.2 | 0.2 | 9.4 | 29.9 | 4.3 |
| Lab All, p_mwu |  | 0 | 0.4646 | 0 | 0 | 0 | 0.0041 | 0.0086 | 0 | 0.3981 | 0.0002 | 0 | 0.005 |
| Lab All, s_mwu |  | 2558.5 | 7547.5 | 1440 | 2389 | 4641 | 6091 | 6204 | 3083 | 7448.5 | 5504 | 3553.5 | 6088.5 |
| Lab All, Std |  | 1.3 | 1.4 | 0.7 | 13 | 18.2 | 9.8 | 24.8 | 16.4 | 13 | 22.7 | 1.4 | 6 |
| Lab All, Iqr |  | 1.9 | 2.3 | 0.8 | 14.7 | 24 | 18.1 | 28 | 18.6 | 15.8 | 27 | 1.7 | 3.1 |
| Lab All, p_dag |  | 0.0258 | 0.0004 | 0 | 0 | 0 | 0.0003 | 0 | 0.0011 | 0 | 0.0003 | 0.0001 | 0 |
| Lab All, p_sha |  | 0.0033 | 0.0149 | 0 | 0 | 0 | 0 | 0 | 0 | 0 | 0 | 0.0006 | 0 |
| Lab All, gaussian |  | False | False | False | False | False | False | False | False | False | False | False | False |
| Lab AHI 0-5, p_tt |  | 0 | 0.805 | 0 | 0 | 0 | 0.8792 | 0.0001 | 0 | 0.4957 | 0 | 0 | 0.0013 |
| Lab AHI 0-5, s_tt |  | 8.3 | 0.2 | 10.1 | 8.1 | -5.7 | 0.2 | 3.9 | -8.5 | 0.7 | -4.5 | -5.4 | 3.3 |
| Lab AHI 0-5, p_medians |  | 0 | 0.4634 | 0 | 0 | 0 | 0 | 0.0001 | 0 | 0.4634 | 0.0001 | 0 | 0.004 |
| Lab AHI 0-5, s_medians |  | 51.2 | 0.5 | 56 | 38 | 25.1 | 18.9 | 15 | 39.9 | 0.5 | 16.2 | 23.3 | 8.3 |
| Lab AHI 0-5, p_mwu |  | 0 | 0.3609 | 0 | 0 | 0 | 0.0024 | 0.0006 | 0 | 0.2011 | 0.0001 | 0 | 0.0009 |
| Lab AHI 0-5, s_mwu |  | 961 | 2666 | 557 | 925 | 1545 | 2043 | 1903 | 850 | 2540 | 1765 | 1297 | 1941.5 |
| Lab AHI 0-5, Std |  | 1.2 | 1.3 | 0.7 | 13.4 | 16.5 | 10.8 | 21.9 | 15.1 | 13 | 21.4 | 1.1 | 3.1 |
| Lab AHI 0-5, Iqr |  | 1.8 | 1.9 | 0.7 | 14.7 | 19.3 | 19.7 | 13.9 | 16.2 | 13.5 | 21.2 | 1.6 | 2.6 |
| Lab AHI 0-5, p_dag |  | 0.435 | 0.8154 | 0.0085 | 0.0001 | 0.001 | 0.0301 | 0 | 0.0013 | 0.0002 | 0.0009 | 0.7701 | 0 |
| Lab AHI 0-5, p_sha |  | 0.2551 | 0.8811 | 0.0004 | 0.0002 | 0 | 0.0001 | 0 | 0.0001 | 0.0001 | 0.0001 | 0.6995 | 0 |
| Lab AHI 0-5, gaussian |  | True | True | False | False | False | False | False | False | False | False | True | False |
| Lab AHI 15-100, p_tt |  | 0 | 0.1552 | 0 | 0 | 0.0014 | 0.1399 | 0.4933 | 0.0003 | 0.0049 | 0.5851 | 0 | 0.9709 |
| Lab AHI 15-100, s_tt |  | 7.8 | 1.4 | 8.3 | 6.2 | -3.3 | 1.5 | -0.7 | -3.7 | 2.9 | -0.5 | -5.2 | 0 |
| Lab AHI 15-100, p_medians |  | 0 | 0.9451 | 0 | 0 | 0.003 | 0.6659 | 0.4269 | 0.0009 | 0.3079 | 0.2471 | 0 | 0.9451 |
| Lab AHI 15-100, s_medians |  | 46.6 | 0 | 57 | 28.9 | 8.8 | 0.2 | 0.6 | 11.1 | 1 | 1.3 | 22.9 | 0 |
| Lab AHI 15-100, p_mwu |  | 0 | 0.1524 | 0 | 0 | 0.0015 | 0.4112 | 0.1524 | 0.0002 | 0.0956 | 0.2423 | 0 | 0.4536 |
| Lab AHI 15-100, s_mwu |  | 624 | 1748 | 435 | 786 | 1348 | 1916 | 1748 | 1231.5 | 1692 | 1815.5 | 963 | 1935.5 |
| Lab AHI 15-100, Std |  | 1.3 | 1.5 | 0.8 | 14.7 | 18.6 | 8.9 | 27.4 | 18.1 | 8.9 | 22.7 | 2 | 10.1 |
| Lab AHI 15-100, Iqr |  | 1.3 | 1.8 | 0.8 | 15.5 | 24.6 | 12.1 | 39.9 | 22.3 | 12.3 | 24 | 2 | 3.9 |
| Lab AHI 15-100, p_dag |  | 0.0595 | 0.4443 | 0 | 0.0048 | 0.0229 | 0.0028 | 0.0999 | 0.9111 | 0.0589 | 0.2116 | 0.0153 | 0 |
| Lab AHI 15-100, p_sha |  | 0.034 | 0.2717 | 0.0005 | 0.0042 | 0.0049 | 0 | 0.0255 | 0.2145 | 0.0009 | 0.0118 | 0.017 | 0 |
| Lab AHI 15-100, gaussian |  | True | True | False | False | False | False | True | True | True | True | False | False |

**Table S5.** Analysis approach A2. Main summary of sleep stage analysis with average HRV and breathing-based sleep stage assessments.

|  | ICU | Sleeplab All | S.Lab AHI<5 | S.Lab AHI>15 |
| --- | --- | --- | --- | --- |
| n subjects | 80 | 190 | 69 | 49 |
| Total Sleep (h) | 8.2 (4.3) | ** 5.5 (1.9) | ** 5.7 (1.8) | ** 5.3 (1.3) |
| Concordant Sleep (h) | 4.0 (2.6) | 4.2 (2.3) | 4.3 (1.9) | 4.0 (1.8) |
| Discordant Sleep (h) | 3.9 (3.7) | ** 1.0 (0.8) | ** 1.0 (0.7) | ** 1.2 (0.8) |
| Discordant Sleep (%) | 41.2 (27.8) | ** 19.4 (14.7) | ** 19.3 (14.7) | ** 22.8 (15.5) |
| Sleep Fraction | 58.2 (24.6) | ** 81.1 (22.3) | ** 84.2 (16.8) | ** 78.4 (19.3) |
| Stage R (%) | 15.1 (26.2) | 14.2 (15.7) | 17.1 (13.9) | 10.6 (17.8) |
| Stage N1 (%) | 23.7 (40.9) | 23.5 (22) | * 19.9 (11.6) | 38.6 (28.2) |
| Stage N2 (%) | 19.5 (16.8) | ** 40.9 (20.3) | ** 45.8 (16.5) | ** 34.9 (19.4) |
| Stage N3 (%) | 17.2 (27.6) | 13.5 (15.5) | 13.9 (13.2) | 10.4 (11.8) |
| Stage N2+N3 (%) | 39.3 (43.5) | * 57.1 (23.5) | ** 60.8 (15.8) | 47 (24.9) |
| SFI | 2.9 (2.0) | * 3.4 (1.6) | 3.1 (1.5) | 3.6 (1.8) |
| Wake Transitions/h | 3.6 (4.7) | 3 (2.5) | * 2.7 (2.0) | 3.9 (2.8) |

**Table S6.** Analysis approach A3. Main summary of sleep stage analysis with average HRV and breathing-based sleep stage assessments.

|  | ICU | Sleeplab All | S.Lab AHI<5 | S.Lab AHI>15 |
| --- | --- | --- | --- | --- |
| n subjects | 80 | 190 | 69 | 49 |
| Total Sleep (h) | 8.2 (4.3) | ** 5.5 (1.9) | ** 5.7 (1.8) | ** 5.3 (1.3) |
| Concordant Sleep (h) | 4.0 (2.6) | 4.2 (2.3) | 4.3 (1.9) | 4.0 (1.8) |
| Discordant Sleep (h) | 3.9 (3.7) | ** 1.0 (0.8) | ** 1.0 (0.7) | ** 1.2 (0.8) |
| Discordant Sleep (%) | 41.2 (27.8) | ** 19.4 (14.7) | ** 19.3 (14.7) | ** 22.8 (15.5) |
| Sleep Fraction | 54.8 (42.4) | ** 81.6 (24.0) | ** 84.1 (19.3) | ** 75.5 (24.6) |
| Stage R (%) | 0.0 (11.9) | ** 7.8 (18.1) | ** 9.3 (19.7) | 1.6 (12.1) |
| Stage N1 (%) | 37.6 (58.1) | * 25.7 (28) | ** 20.9 (13.9) | 47.5 (39.9) |
| Stage N2 (%) | 19.3 (25.7) | ** 43.6 (18.6) | ** 47.9 (16.2) | ** 35.1 (22.3) |
| Stage N3 (%) | 11.1 (31.3) | 12.5 (15.8) | 14.9 (13.5) | 7.6 (12.3) |
| Stage N2+N3 (%) | 33.6 (63.5) | * 58.7 (27) | ** 64.5 (21.2) | 46.1 (24.0) |
| SFI | 1.3 (2.0) | ** 2.9 (1.7) | ** 2.8 (1.6) | ** 3.0 (2.0) |
| Wake Transitions/h | 4.1 (6.6) | * 2.6 (3.1) | ** 2.1 (2.6) | 4.0 (3.9) |

**Table S7.**
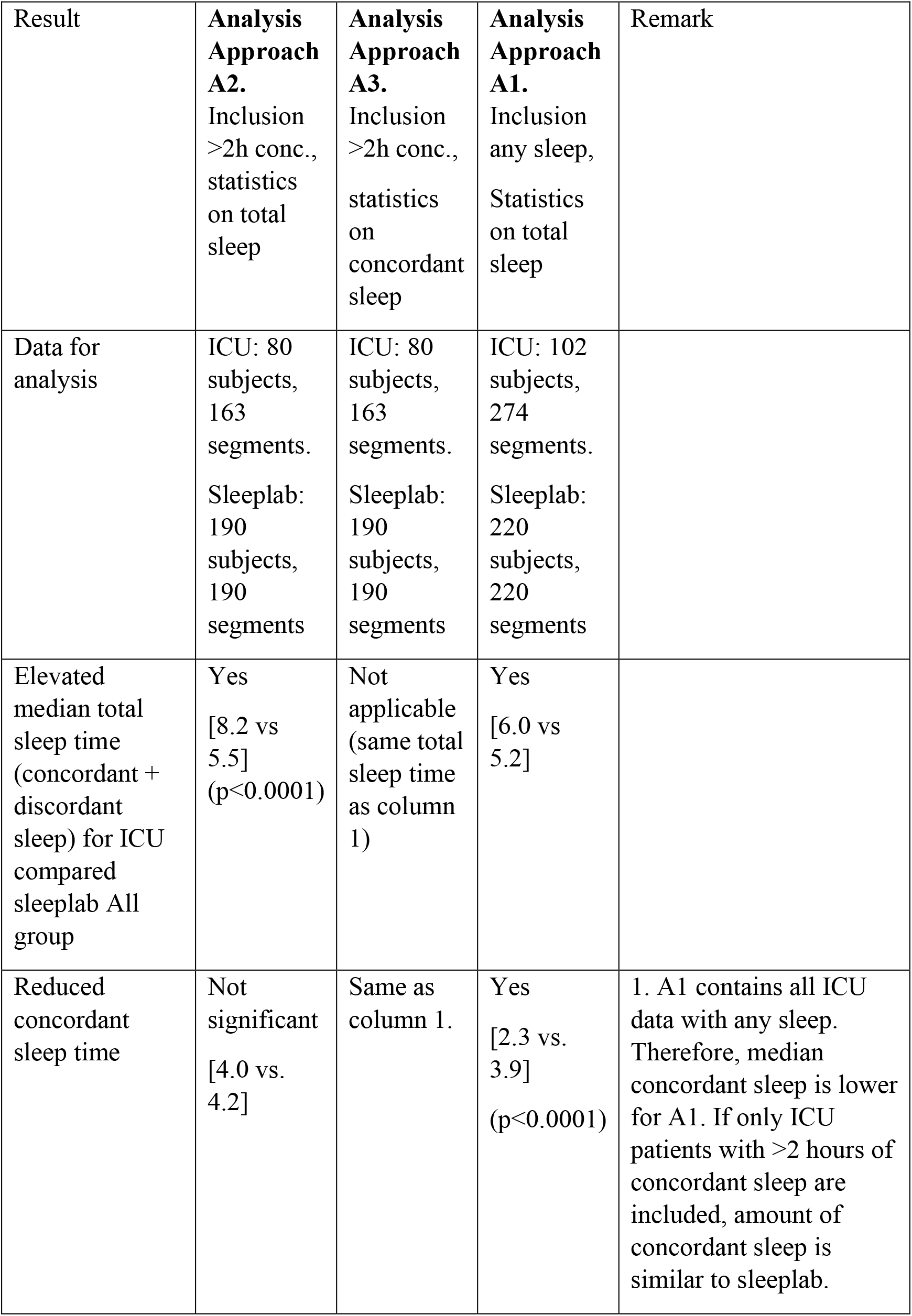
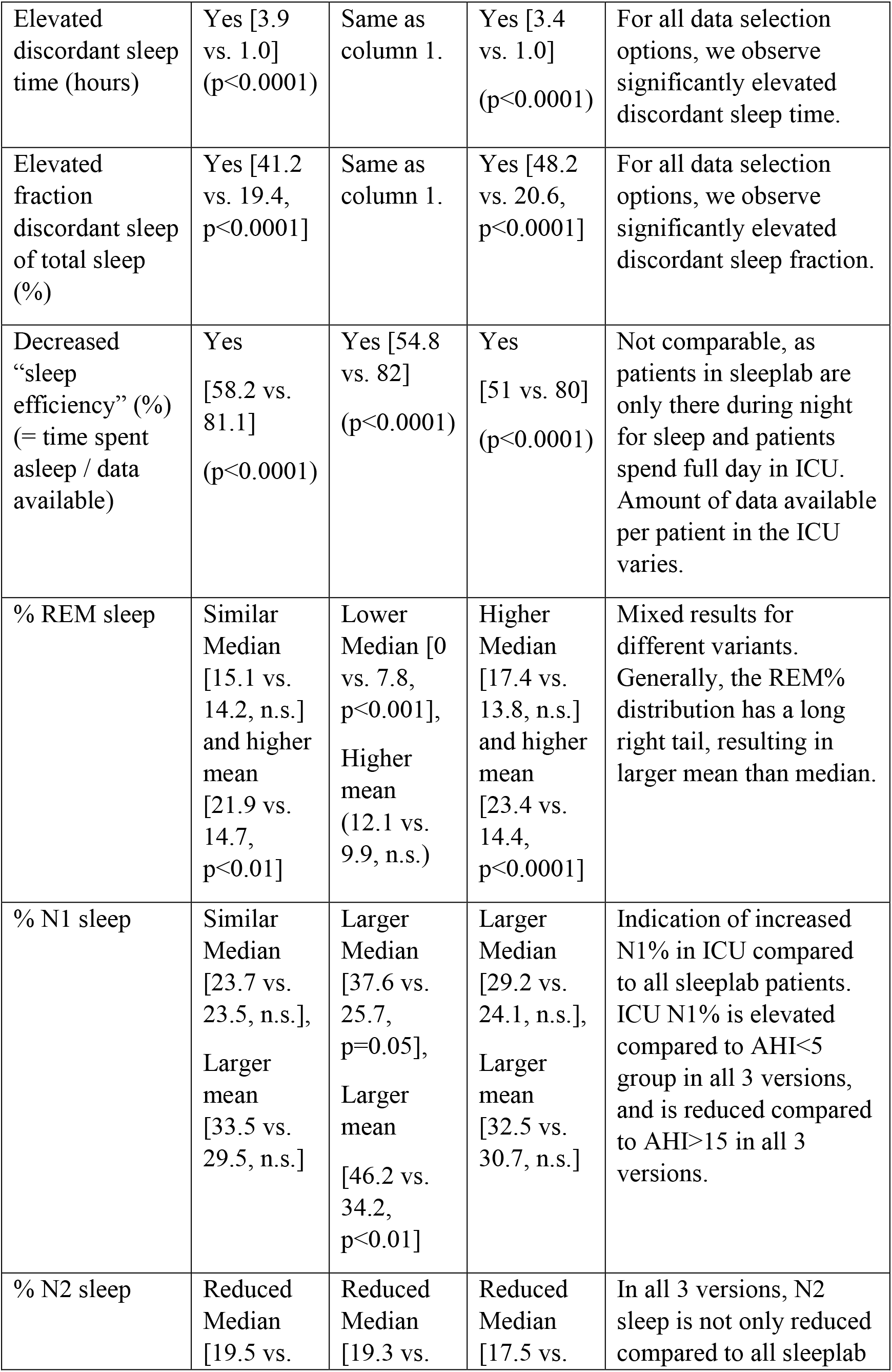
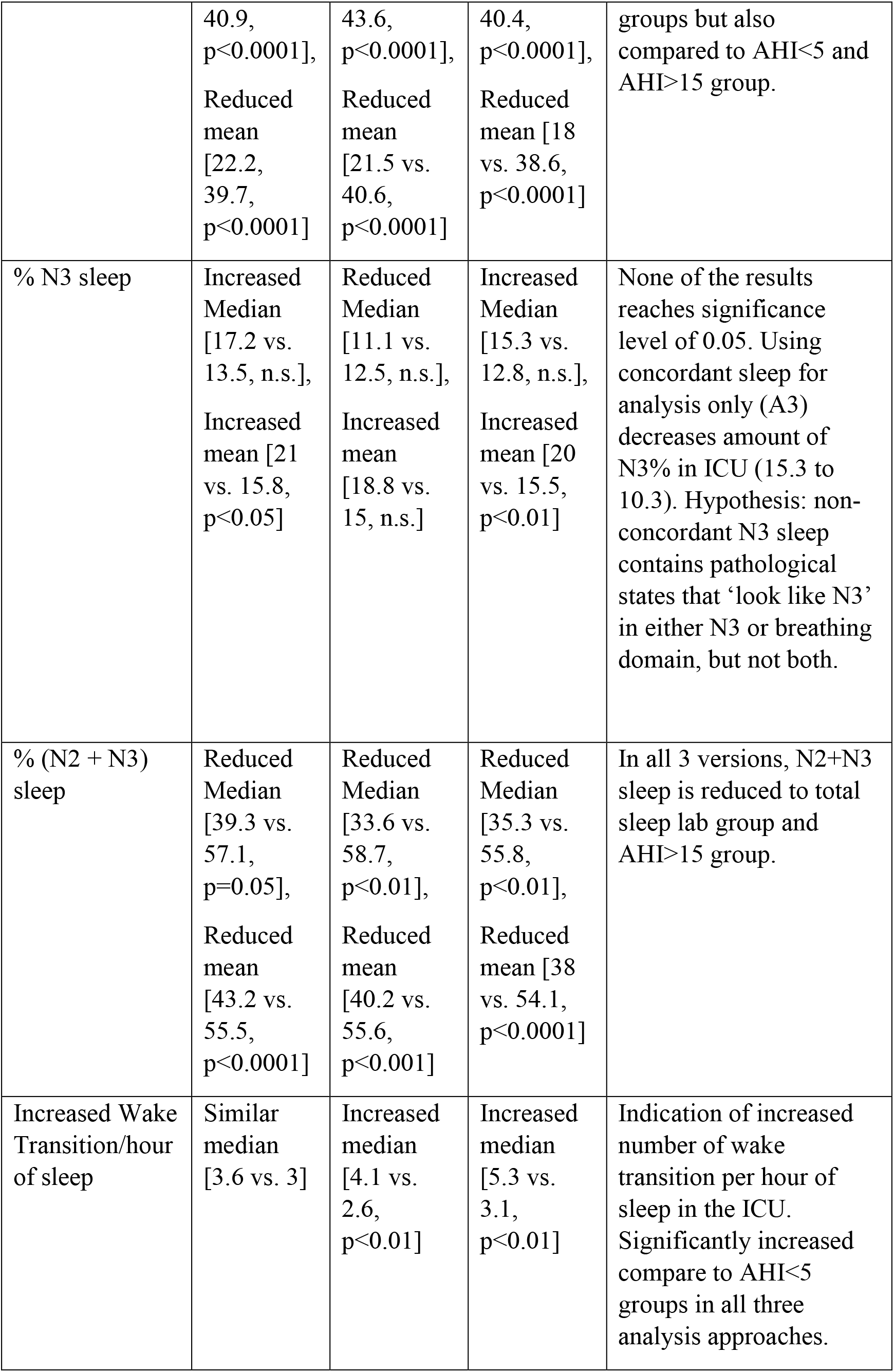
Summary and remarks interpretation for sleep stage analysis for ICU and sleeplab comparison.

| Result | **Analysis Approach A2.**<br>Inclusion >2h conc., statistics on total sleep | **Analysis Approach A3.**<br>Inclusion >2h conc., statistics on concordant sleep | **Analysis Approach A1.**<br>Inclusion any sleep, Statistics on total sleep | Remark |
| --- | --- | --- | --- | --- |
| Data for analysis | ICU: 80 subjects, 163 segments.<br>Sleeplab: 190 subjects, 190 segments | ICU: 80 subjects, 163 segments.<br>Sleeplab: 190 subjects, 190 segments | ICU: 102 subjects, 274 segments.<br>Sleeplab: 220 subjects, 220 segments |  |
| Elevated median total sleep time (concordant + discordant sleep) for ICU compared sleeplab All group | Yes<br>[8.2 vs 5.5]<br>($p < 0.0001$) | Not applicable (same total sleep time as column 1) | Yes<br>[6.0 vs 5.2] | |
| Reduced concordant sleep time | Not significant<br>[4.0 vs. 4.2] | Same as column 1. | Yes<br>[2.3 vs. 3.9]<br>($p < 0.0001$) | 1. A1 contains all ICU data with any sleep. Therefore, median concordant sleep is lower for A1. If only ICU patients with >2 hours of concordant sleep are included, amount of concordant sleep is similar to sleeplab. |
| Elevated discordant sleep time (hours) | Yes [3.9 vs. 1.0] (p<0.0001) | Same as column 1. | Yes [3.4 vs. 1.0] (p<0.0001) | For all data selection options, we observe significantly elevated discordant sleep time. |
| Elevated fraction discordant sleep of total sleep (%) | Yes [41.2 vs. 19.4, p<0.0001] | Same as column 1. | Yes [48.2 vs. 20.6, p<0.0001] | For all data selection options, we observe significantly elevated discordant sleep fraction. |
| Decreased “sleep efficiency” (%) (= time spent asleep / data available) | Yes [58.2 vs. 81.1] (p<0.0001) | Yes [54.8 vs. 82] (p<0.0001) | Yes [51 vs. 80] (p<0.0001) | Not comparable, as patients in sleep lab are only there during night for sleep and patients spend full day in ICU. Amount of data available per patient in the ICU varies. |
| % REM sleep | Similar Median [15.1 vs. 14.2, n.s.] and higher mean [21.9 vs. 14.7, p<0.01] | Lower Median [0 vs. 7.8, p<0.001], Higher mean (12.1 vs. 9.9, n.s.) | Higher Median [17.4 vs. 13.8, n.s.] and higher mean [23.4 vs. 14.4, p<0.0001] | Mixed results for different variants. Generally, the REM% distribution has a long right tail, resulting in larger mean than median. |
| % N1 sleep | Similar Median [23.7 vs. 23.5, n.s.], Larger mean [33.5 vs. 29.5, n.s.] | Larger Median [37.6 vs. 25.7, p=0.05], Larger mean [46.2 vs. 34.2, p<0.01] | Larger Median [29.2 vs. 24.1, n.s.], Larger mean [32.5 vs. 30.7, n.s.] | Indication of increased N1% in ICU compared to all sleep lab patients. ICU N1% is elevated compared to AHI<5 group in all 3 versions, and is reduced compared to AHI>15 in all 3 versions. |
| % N2 sleep | Reduced Median [19.5 vs. | Reduced Median [19.3 vs. | Reduced Median [17.5 vs. | In all 3 versions, N2 sleep is not only reduced compared to all sleep lab |
|  | 40.9,<br>p<0.0001],<br>Reduced<br>mean<br>[22.2,<br>39.7,<br>p<0.0001] | 43.6,<br>p<0.0001],<br>Reduced<br>mean<br>[21.5 vs.<br>40.6,<br>p<0.0001] | 40.4,<br>p<0.0001],<br>Reduced<br>mean [18<br>vs. 38.6,<br>p<0.0001] | groups but also<br>compared to AHI<5 and<br>AHI>15 group. |
| % N3 sleep | Increased<br>Median<br>[17.2 vs.<br>13.5, n.s.],<br>Increased<br>mean [21<br>vs. 15.8,<br>p<0.05] | Reduced<br>Median<br>[11.1 vs.<br>12.5, n.s.],<br>Increased<br>mean<br>[18.8 vs.<br>15, n.s.] | Increased<br>Median<br>[15.3 vs.<br>12.8, n.s.],<br>Increased<br>mean [20<br>vs. 15.5,<br>p<0.01] | None of the results<br>reaches significance<br>level of 0.05. Using<br>concordant sleep for<br>analysis only (A3)<br>decreases amount of<br>N3% in ICU (15.3 to<br>10.3). Hypothesis: non-<br>concordant N3 sleep<br>contains pathological<br>states that ‘look like N3’<br>in either N3 or breathing<br>domain, but not both. |
| % (N2 + N3)<br>sleep | Reduced<br>Median<br>[39.3 vs.<br>57.1,<br>p=0.05],<br>Reduced<br>mean<br>[43.2 vs.<br>55.5,<br>p<0.0001] | Reduced<br>Median<br>[33.6 vs.<br>58.7,<br>p<0.01],<br>Reduced<br>mean<br>[40.2 vs.<br>55.6,<br>p<0.001] | Reduced<br>Median<br>[35.3 vs.<br>55.8,<br>p<0.01],<br>Reduced<br>mean [38<br>vs. 54.1,<br>p<0.0001] | In all 3 versions, N2+N3<br>sleep is reduced to total<br>sleep lab group and<br>AHI>15 group. |
| Increased Wake<br>Transition/hour<br>of sleep | Similar<br>median<br>[3.6 vs. 3] | Increased<br>median<br>[4.1 vs.<br>2.6,<br>p<0.01] | Increased<br>median<br>[5.3 vs.<br>3.1,<br>p<0.01] | Indication of increased<br>number of wake<br>transition per hour of<br>sleep in the ICU.<br>Significantly increased<br>compare to AHI<5<br>groups in all three<br>analysis approaches. |

#### Latent feature representation of sleep

The last hidden layers’ dimensions (D_LHL) for the Long Short-Term Memory (LSTM) networks were (40, 1) for the HRV-based model and (200, 1) for the breathing-based model. We performed two UMAPs (Uniform Manifold Approximation and Projection) separately for the data resulting from the HRV-based and breathing-based models. The python package UMAP(3) was used to perform the UMAPs, and python packages matplotlib and seaborn were used for visualization.

Pseudo code:

1. Select either HRV-based or breathing-based model, and concatenate all last hidden layer activations for both ICU and sleeplab data, resulting in an array (‘LHL array’) with shape: (N_ALL, D_LHL) with N_ALL = 693,401, number of epochs from sleeplab data (324, 928) plus number of epochs from ICU data (368, 473), and D_LHL=40 for HRV- and D_LHL=200 for breathing-based model.
2. Perform unsupervised UMAP with input LHL array and select the first two dimensions of the resulting UMAP embedding, resulting in an array (N_ALL, 2). Parameters: number of neighbors=15, minimum distance=0.1, metric = “euclidean”.
3. For **Figure 3A**, plot UMAP embeddings for sleeplab data only, ICU data only, and sleeplab and ICU data combined (three figure panel columns). For each plot, color the data according to the assigned sleep stage.
4. For **Figure 3B**, for each panel column, select a sleep stage (W, R, N1, N2, N3) and estimate kernel densities for the UMAP embeddings of both the sleeplab and ICU data. Plot the 10% iso-proportion levels.

#### Disagreement HRV and breathing model - error analysis

In order to identify features in the cardiovascular and pulmonary system that are associated with discordant sleep, we compute breathing and heart rate variability related features and compare the values of those features for agreeing and discordant sleep. We use the four breathing features as described above: respiratory rate, inter-breath-intervals, variability index, and ventilation CVar. For HRV, we compute a root mean squared successive difference of NN intervals (RMSSD), very low frequency (VLF, 0.0033-0.04 Hz), low frequency (0.04-0.15 Hz), and high frequency (HF, 0.15-0.4 Hz) with a 5-minute sliding window, using Lomb-Scargle periodogram for spectral analysis. We further compute Cardiopulmonary Coupling (CPC)(4), a frequency domain analysis method that combines cross spectral power and coherence of the respiratory and HRV signals. Unlike in the original publication where respiratory signal is estimated from the ECG signal(4), we use the wearable respiratory signal. From the CPC spectrogram, we compute the amounts of low frequency coupling (LFC, 0.01-0.1 Hz) and high frequency coupling (HFC, 0.1-0.4 Hz) with an 8.5-minute sliding window.

Summary features computed:

a. Heart Rate Variability: NN interval, mean very low frequency power (VLF, 0.0033-0.04 Hz), low frequency power (0.04-0.15 Hz), high frequency power (HF, 0.15-0.4 Hz), RMSSD of NN intervals.
b. Breathing: inter-breath-interval, respiratory rate, variability index, ventilation coefficient of variation.
c. Cardiopulmonary Coupling: low frequency coupling (LFC, 0.01-0.1 Hz), high frequency coupling (HFC, 0.1-0.4 Hz).

We computed the mean feature values (e.g. mean NN interval) for concordant and discordant parts for every 24-hour segment (inclusion criteria: at least 2 hours of concordant sleep) and performed a Wilcoxon signed-rank test with an alpha value of 0.01 for each feature pair. Features were computed for each sleep stage. Note: Discordant sleep is partially asymmetric, an epoch that is classified as N2 by model A and as Wake by model B is included for model A analysis here but not for model B analysis. Further, if model A classifies an epoch as N1, and model B as N3, the epoch is treated as ‘N1-discordant’ for model A and ‘N3-discordant’ for model B.

**Table S8.** Median and interquartile ranges for sleep indices for ICU patients grouped by their primary or main condition. Data is graphically displayed in Figure 6.

|  | N | Total Sleep (h) | Concordant (h) | Discordant (h) | Discordant (%) | Sleep (%) | R (%) | N1 (%) | N2 (%) | N3 (%) | N2+N3 (%) | Wake Trans. |
| --- | --- | --- | --- | --- | --- | --- | --- | --- | --- | --- | --- | --- |
| Sepsis | 13 | 6.2 | 3.5 | 2.8 | 0.38 | 0.42 | 0.04 | 0.33 | 0.01 | 0.41 | 0.46 | 7.9 |
|  |  | [4.4, 7.5] | [2.1, 4.1] | [1.0, 3.8] | [0.17, 0.5] | [0.22, 0.53] | [0.0, 0.13] | [0.16, 0.82] | [0.0, 0.06] | [0.06, 0.6] | [0.15, 0.72] | [4.1, 11.9] |
| Shock | 23 | 6 | 3.7 | 1.7 | 0.31 | 0.37 | 0.04 | 0.28 | 0.05 | 0.3 | 0.3 | 7 |
|  |  | [3.2, 7.5] | [1.8, 4.7] | [0.6, 3.8] | [0.14, 0.45] | [0.25, 0.49] | [0.0, 0.2] | [0.07, 0.77] | [0.0, 0.13] | [0.05, 0.51] | [0.07, 0.75] | [2.6, 10.9] |
| Heart Failure | 8 | 6.7 | 3.2 | 2 | 0.43 | 0.41 | 0.04 | 0.26 | 0.01 | 0.59 | 0.63 | 6.4 |
|  |  | [1.3, 8.3] | [1.1, 5.0] | [0.3, 3.8] | [0.08, 0.61] | [0.07, 0.52] | [0.01, 0.08] | [0.04, 0.36] | [0.0, 0.04] | [0.32, 0.89] | [0.36, 0.91] | [2.1, 26.2] |
| Encephalopathy | 9 | 7.4 | 3.7 | 2.3 | 0.54 | 0.45 | 0.01 | 0.33 | 0.04 | 0.17 | 0.18 | 4.8 |
|  |  | [5.0, 7.6] | [2.7, 4.1] | [0.6, 4.7] | [0.17, 0.62] | [0.25, 0.53] | [0.0, 0.13] | [0.17, 0.82] | [0.02, 0.07] | [0.05, 0.48] | [0.08, 0.7] | [3.5, 19.0] |
| Anemia | 15 | 5.9 | 3.3 | 2.3 | 0.36 | 0.32 | 0 | 0.31 | 0.03 | 0.37 | 0.54 | 5.8 |
|  |  | [4.4, 7.3] | [2.7, 4.5] | [1.3, 3.1] | [0.2, 0.46] | [0.27, 0.51] | [0.0, 0.06] | [0.12, 0.78] | [0.01, 0.13] | [0.06, 0.72] | [0.15, 0.84] | [3.0, 12.9] |
| Hemorrhage | 8 | 7.3 | 4.2 | 3.2 | 0.32 | 0.42 | 0.34 | 0.36 | 0.03 | 0.06 | 0.1 | 6.4 |
|  |  | [4.7, 9.5] | [3.2, 6.4] | [0.5, 4.1] | [0.12, 0.42] | [0.39, 0.69] | [0.04, 0.7] | [0.18, 0.6] | [0.0, 0.1] | [0.01, 0.18] | [0.05, 0.39] | [3.9, 9.3] |
| Pneumonia | 13 | 6.1 | 2.7 | 3.1 | 0.49 | 0.37 | 0.07 | 0.31 | 0.06 | 0.17 | 0.3 | 5.8 |
|  |  | [4.9, 7.5] | [1.5, 3.8] | [2.3, 4.5] | [0.28, 0.59] | [0.25, 0.53] | [0.0, 0.26] | [0.24, 0.81] | [0.01, 0.09] | [0.06, 0.41] | [0.15, 0.52] | [4.8, 10.7] |
| Resp. Failure | 14 | 5.5 | 2.6 | 2.3 | 0.51 | 0.37 | 0.1 | 0.18 | 0.02 | 0.23 | 0.31 | 4.1 |
|  |  | [3.6, 9.0] | [1.5, 4.3] | [0.8, 5.3] | [0.25, 0.59] | [0.25, 0.54] | [0.0, 0.43] | [0.04, 0.31] | [0.0, 0.08] | [0.07, 0.58] | [0.18, 0.6] | [2.0, 7.3] |
| AKI | 24 | 6.6 | 3.7 | 2.8 | 0.35 | 0.43 | 0.06 | 0.22 | 0.02 | 0.44 | 0.5 | 4.4 |
|  |  | [4.6, 8.7] | [2.1, 5.2] | [0.6, 3.8] | [0.12, 0.54] | [0.32, 0.64] | [0.0, 0.22] | [0.08, 0.46] | [0.0, 0.08] | [0.1, 0.59] | [0.11, 0.73] | [3.2, 9.0] |
| Pneumothorax | 10 | 6.9 | 3.5 | 3.6 | 0.53 | 0.42 | 0.19 | 0.15 | 0.03 | 0.57 | 0.62 | 4.4 |
|  |  | [6.0, 7.9] | [2.7, 4.4] | [1.5, 5.3] | [0.41, 0.63] | [0.38, 0.53] | [0.08, 0.25] | [0.08, 0.28] | [0.01, 0.12] | [0.42, 0.62] | [0.49, 0.7] | [3.6, 5.6] |
| COPD | 6 | 7.8 | 4.9 | 2.6 | 0.36 | 0.49 | 0.05 | 0.09 | 0.03 | 0.86 | 0.87 | 3.2 |
|  |  | [5.5, 10.0] | [3.0, 6.4] | [1.3, 3.5] | [0.35, 0.45] | [0.32, 0.57] | [0.02, 0.09] | [0.04, 0.15] | [0.01, 0.14] | [0.64, 0.91] | [0.77, 0.94] | [1.4, 5.2] |
| Other Medical | 3 | 6.4 | 3.4 | 4.8 | 0.75 | 0.62 | 0.04 | 0.09 | 0.01 | 0.91 | 0.91 | 3 |
|  |  | [6.4, 10.7] | [2.5, 3.9] | [3.4, 8.3] | [0.53, 0.75] | [0.54, 0.71] | [0.02, 0.31] | [0.06, 0.19] | [0.01, 0.02] | [0.51, 0.91] | [0.52, 0.92] | [2.3, 4.9] |
| GI Hernia | 7 | 7 | 3.2 | 3.7 | 0.5 | 0.39 | 0.11 | 0.17 | 0.05 | 0.6 | 0.7 | 4.1 |
|  |  | [5.0, 7.5] | [2.8, 3.7] | [2.6, 4.2] | [0.45, 0.55] | [0.34, 0.46] | [0.03, 0.23] | [0.13, 0.25] | [0.02, 0.06] | [0.42, 0.68] | [0.54, 0.74] | [3.6, 5.9] |
| Liver | 5 | 4.8 | 2.7 | 1.7 | 0.54 | 0.32 | 0.34 | 0.16 | 0.14 | 0.09 | 0.43 | 4.7 |
|  |  | [3.9, 6.2] | [2.6, 3.3] | [1.2, 3.5] | [0.31, 0.54] | [0.26, 0.32] | [0.3, 0.63] | [0.15, 0.19] | [0.01, 0.33] | [0.08, 0.12] | [0.22, 0.5] | [4.6, 8.2] |
| Other Surgical | 43 | 6.8 | 4.1 | 2.3 | 0.34 | 0.5 | 0.12 | 0.17 | 0.11 | 0.24 | 0.45 | 4.3 |
|  |  | [4.9, 9.2] | [2.8, 5.7] | [1.1, 3.7] | [0.18, 0.48] | [0.3, 0.66] | [0.02, 0.34] | [0.06, 0.41] | [0.01, 0.21] | [0.05, 0.55] | [0.18, 0.77] | [2.1, 7.2] |
| Kruskal statistic, p-value |  | 7.7, 0.904 | 12.5, 0.565 | 8.0, 0.888 | 16.6, 0.278 | 10.8, 0.705 | 18.4, 0.191 | 16.6, 0.279 | 15.7, 0.33 | 21.9, 0.081 | 18.4, 0.189 | 11.6, 0.64 |

**Table S9.** Error Analysis – Details. HRV and respiratory features (rows in table) were computed for concordant sleep and discordant sleep, for both the HRV-based and breathing-based sleep staging models, and for each sleep stage. Columns contain number of 24-hour segments available after inclusion criteria (minimum 2 hours of concordant sleep), feature values, and the Mann–Whitney U test (both test statistic and p-value). The ‘significance’ column shows ‘True’ if the p-value is less than 0.01 for any of the HRV- or breathing-based sleep staging models.

|  | Breathing |  |  |  | HRV |  |  |  | Signif. |
| --- | --- | --- | --- | --- | --- | --- | --- | --- | --- |
|  | N | Conc. | Disc. | test | N | Conc. | Disc. | test |  |
| N1, hrv_nn | 100 | 0.756 | 0.76 | 2091, 0.13564 | 71 | 0.791 | 0.792 | 1229, 0.77889 | False |
| N1, hrv_vlf | 80 | 1914.6 | 1474.2 | 840, 0.00018 | 59 | 2297.9 | 2172.6 | 773, 0.3979 | True |
| N1, hrv_lf | 80 | 1129.3 | 914.3 | 916, 0.00073 | 59 | 1376.2 | 1388.9 | 848, 0.78003 | True |
| N1, hrv_hf | 80 | 2055 | 1793.7 | 994, 0.00268 | 59 | 2858.1 | 3010.8 | 847, 0.77425 | True |
| N1, hrv_rmssd | 80 | 65.5 | 60 | 926, 0.00087 | 59 | 76.3 | 73.1 | 655, 0.08256 | True |
| N1, ibi | 104 | 3.98 | 4.01 | 2595, 0.66155 | 71 | 3.95 | 3.55 | 518, 1e-05 | True |
| N1, rr_10s_smooth | 104 | 17.7 | 17.4 | 2246, 0.11653 | 71 | 17.8 | 19.5 | 537, 2e-05 | True |
| N1, instability_30sec | 104 | 0.243 | 0.238 | 2286, 0.14992 | 71 | 0.225 | 0.194 | 653, 0.00034 | True |
| N1, ventilation_cvar_30sec | 104 | 0.235 | 0.233 | 2446, 0.35707 | 71 | 0.218 | 0.181 | 554, 3e-05 | True |
| N1, cpc_lfc | 80 | 4.29 | 3.38 | 978, 0.00208 | 59 | 4.92 | 2.67 | 588, 0.02498 | True |
| N1, cpc_hfc | 80 | 0.408 | 0.348 | 927, 0.00089 | 59 | 0.415 | 0.338 | 715, 0.19944 | True |
| N2, hrv_nn | 109 | 0.786 | 0.769 | 2019, 0.00309 | 35 | 0.867 | 0.853 | 229, 0.15895 | True |
| N2, hrv_vlf | 89 | 1258.7 | 1784.9 | 1008, 5e-05 | 35 | 1055.9 | 1122.8 | 294, 0.73087 | True |
| N2, hrv_lf | 89 | 910.9 | 1200.5 | 1170, 0.00066 | 35 | 726.7 | 726.9 | 309, 0.92171 | True |
| N2, hrv_hf | 89 | 2527 | 2355.4 | 1332, 0.00608 | 35 | 2272 | 2377.2 | 300, 0.80592 | True |
| N2, hrv_rmssd | 89 | 67 | 70.2 | 1378, 0.01062 | 35 | 55.3 | 54.6 | 311, 0.94776 | False |
| N2, ibi | 112 | 3.75 | 3.59 | 1677, 2e-05 | 35 | 4.02 | 3.82 | 162, 0.01221 | True |
| N2, rr_10s_smooth | 112 | 18.1 | 18.8 | 1485, 0.0 | 35 | 16.8 | 17.9 | 121, 0.00149 | True |
| N2, instability_30sec | 112 | 0.185 | 0.167 | 2121, 0.00246 | 35 | 0.182 | 0.183 | 284, 0.61163 | True |
| N2, ventilation_cvar_30sec | 112 | 0.184 | 0.162 | 2016, 0.00086 | 35 | 0.179 | 0.167 | 311, 0.94776 | True |
| N2, cpc_lfc | 89 | 2.11 | 1.88 | 1496, 0.03824 | 35 | 2.78 | 1.39 | 302, 0.83138 | False |
| N2, cpc_hfc | 89 | 1.07 | 1.38 | 1939, 0.79502 | 35 | 1.025 | 0.887 | 312, 0.96081 | False |
| N3, hrv_nn | 30 | 0.823 | 0.787 | 119, 0.01957 | 74 | 0.8 | 0.771 | 695, 0.00019 | True |
| N3, hrv_vlf | 25 | 618 | 1793.2 | 21, 3e-05 | 66 | 587.8 | 765.5 | 899, 0.18712 | True |
| N3, hrv_lf | 25 | 733.4 | 1366.3 | 50, 0.00163 | 66 | 618.7 | 652.8 | 1021, 0.58934 | True |
| N3, hrv_hf | 25 | 2252.3 | 2989.3 | 62, 0.00558 | 66 | 1586.7 | 1574.8 | 961, 0.35597 | True |
| N3, hrv_rmssd | 25 | 69.2 | 78.8 | 64, 0.00673 | 66 | 56.6 | 56.9 | 990, 0.46062 | True |
| N3, ibi | 31 | 3.26 | 3.16 | 193, 0.28112 | 74 | 3.66 | 3.68 | 1343, 0.81054 | False |
| N3, rr_10s_smooth | 31 | 20.2 | 20.7 | 182, 0.19588 | 74 | 18.2 | 18.5 | 1105, 0.12804 | False |
| N3, instability_30sec | 31 | 0.134 | 0.124 | 235, 0.79891 | 74 | 0.146 | 0.179 | 485, 0.0 | True |
| N3, ventilation_cvar_30sec | 31 | 0.131 | 0.117 | 182, 0.19588 | 74 | 0.143 | 0.166 | 634, 5e-05 | True |
| N3, cpc_lfc | 25 | 0.193 | 1.122 | 17, 1e-05 | 66 | 0.82 | 1.341 | 528, 0.00023 | True |
| N3, cpc_hfc | 25 | 0.344 | 0.474 | 94, 0.0667 | 66 | 0.842 | 0.52 | 1022, 0.59375 | False |
| R, hrv_nn | 50 | 0.771 | 0.762 | 493, 0.16305 | 41 | 0.767 | 0.766 | 386, 0.56418 | False |
| R, hrv_vlf | 44 | 1899 | 1922.2 | 475, 0.81545 | 40 | 1948.2 | 2194.9 | 384, 0.72673 | False |
| R, hrv_lf | 44 | 1090.4 | 1137.7 | 390, 0.22044 | 40 | 970.3 | 1138 | 332, 0.29444 | False |
| R, hrv_hf | 44 | 1803 | 2048.1 | 291, 0.01728 | 40 | 1207 | 1408.9 | 289, 0.10387 | False |
| R, hrv_rmssd | 44 | 58.2 | 64.2 | 248, 0.00394 | 40 | 44.3 | 47.4 | 303, 0.15037 | True |
| R, ibi | 53 | 3.54 | 3.57 | 711, 0.96822 | 41 | 3.57 | 3.72 | 278, 0.04814 | False |
| R, rr_10s_smooth | 53 | 19.1 | 19 | 688, 0.80766 | 41 | 18.9 | 18.2 | 253, 0.02144 | False |
| R, instability_30sec | 53 | 0.198 | 0.188 | 487, 0.04309 | 41 | 0.2 | 0.196 | 371, 0.44069 | False |
| R, ventilation_cvar_30sec | 53 | 0.179 | 0.167 | 459, 0.02316 | 41 | 0.178 | 0.183 | 375, 0.47202 | False |
| R, cpc_lfc | 44 | 4.1 | 3.1 | 443, 0.54395 | 40 | 3.48 | 3.88 | 354, 0.45162 | False |
| R, cpc_hfc | 44 | 0.445 | 0.617 | 253, 0.00474 | 40 | 0.34 | 0.614 | 297, 0.1288 | True |

**Table S10.** Shows the significant effect directions of Table S9 for easier readability. I.e. ‘+’/’-‘ indicate a significant (0.01 level) increase/decrease of a feature in discordant sleep compared to concordant sleep.

|  | Breathing, Effect<br>Discordant Sleep | HRV, Effect<br>Discordant Sleep |
| --- | --- | --- |
| N1, hrv_vlf | - |  |
| N1, hrv_lf | - |  |
| N1, hrv_hf | - |  |
| N1, hrv_rmssd | - |  |
| N1, ibi |  | - |
| N1, rr_10s_smooth |  | + |
| N1, instability_30sec |  | - |
| N1, ventilation_cvar_30sec |  | - |
| N1, cpc_lfc | - |  |
| N1, cpc_hfc | - |  |
| N2, hrv_nn | - |  |
| N2, hrv_vlf | + |  |
| N2, hrv_lf | + |  |
| N2, ibi | - |  |
| N2, rr_10s_smooth | + | + |
| N2, instability_30sec | - |  |
| N2, ventilation_cvar_30sec | - |  |
| N3, hrv_nn |  | - |
| N3, hrv_vlf | + |  |
| N3, hrv_lf | + |  |
| N3, hrv_hf | + |  |
| N3, hrv_rmssd | + |  |
| N3, instability_30sec |  | + |
| N3, ventilation_cvar_30sec |  | + |
| N3, cpc_lfc | + | + |
| R, hrv_rmssd | + |  |
| R, cpc_hfc | + |  |

For each 24-hour segment included (inclusion criteria: at least 2 hours of concordant sleep), we obtained the proportion of discordant sleep, which is a number in the [0, 1] interval. To make the variable unbounded and more gaussian, we used the following transformation:

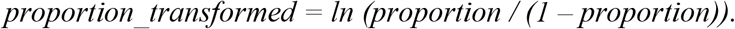

We computed the mean, standard deviation, 0.25-, 0.50-, and 0.75 quantiles for each feature for every 24-hour segment and for each sleep stage. This lead to 886 features, and as a consequence, we used the LASSO penalty for multilinear regression. We start with a penalty of 0 and with a step size of 0.1, we increase the penalty until 10. For each penalty value, we train a LASSO model, obtain the variables included in the model and with those variables train a vanilla multilinear regression model for which we obtain the F-test statistic, F-test p-value and the r-squared. The result is shown in **Figure S4**, indicating overfitted models close to a penalty of 0 and underfitted models close to a penalty of 10. A penalty of 4 results in a regression model that contains 53 variables, an F-statistic of 1.51, a F-test p-value of 0.036, and an r-squared of 0.42. Hence, we obtained evidence that up to 42% of the variance in the discordance sleep proportion over a full 24-hour segment can be explained by HRV and breathing based features that are computed over pooled discordant and concordant parts.

**Figure S5.**
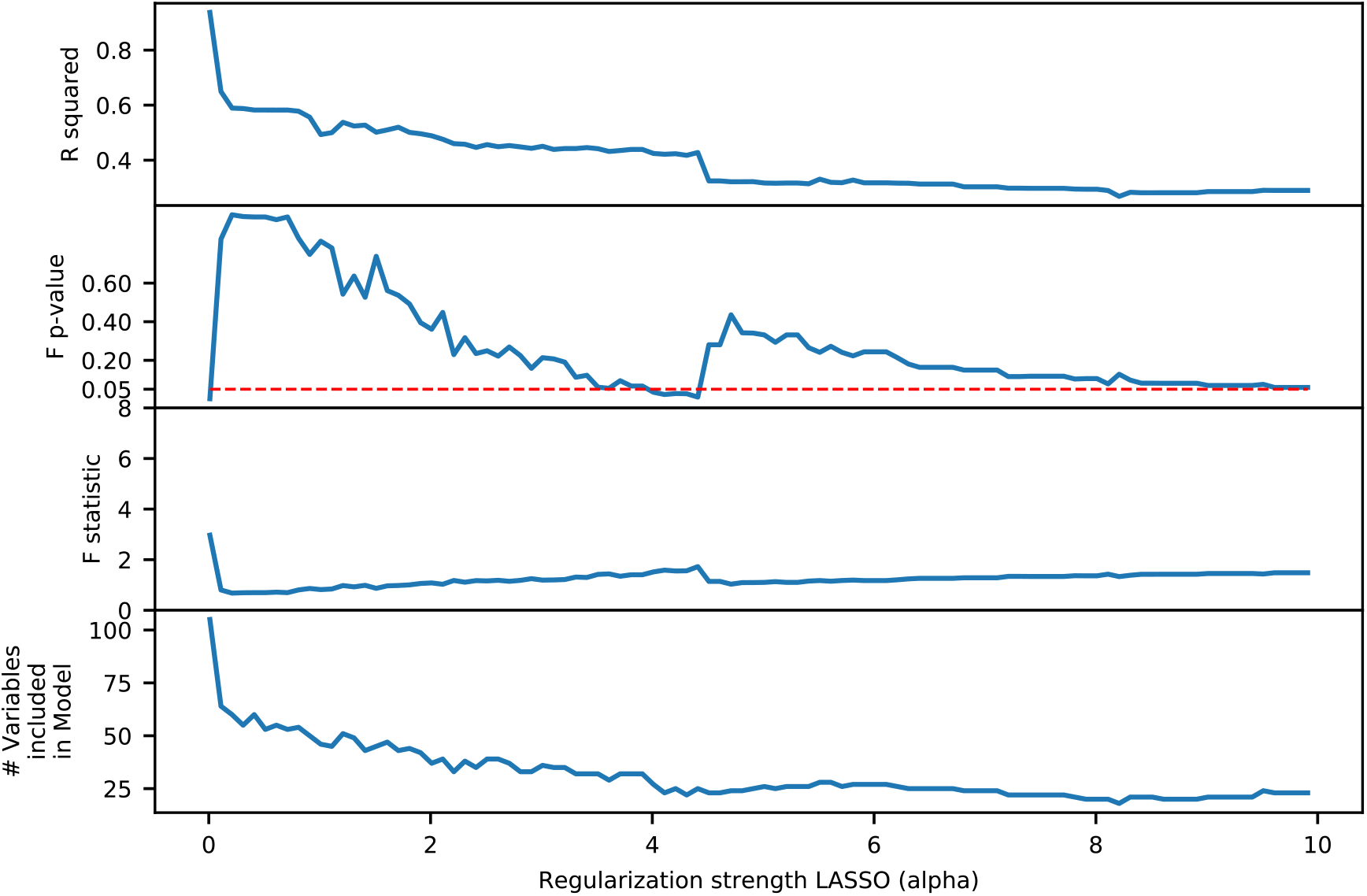
Model development for error analysis. We trained linear regression models with varying LASSO penalty. A penalty of 4 results in a regression model that contains 53 variables, an F-statistic of 1.51, a F-test p-value of 0.036, and an r-squared of 0.42, showing 42% of the variance in discordant sleep proportion is explained by this model.

**Table S11.** Correlations between proportion of discordant sleep and Sequential Organ Failure Assessment (SOFA) score or medications administered. Medications included in the analysis: Opioids (Buprenorphine, Morphine, Oxycodone, Hydrocodone, Hydromorphone, Fentanyl, Meperidine, Codeine, Tramadol), Benzodiazepines (Alprazolam, Chlordiazepoxide, Clonazepam, Diazepam, Lorazepam, Midazolam, Oxazepam, Phenobarbital, Propofol), Antipsychotics (Olanzapine, Clozapine, Thiothixene, Haloperidol, Fluphenazine, Prochlorperazine, Trifluoperazine, Loxapine, Quetiapine, Asenapine). Medications within a category were converted to equivalent doses before summed(5–7).

|  | Pearson<br>Correlation | Pearson<br>p-value | Spearman<br>Correlation | Spearman<br>p-value |
| --- | --- | --- | --- | --- |
| SOFA | 0.059 | 0.457 | 0.090 | 0.256 |
| Opioids | -0.086 | 0.276 | -0.080 | 0.308 |
| Benzodiazepines | -0.022 | 0.776 | 0.012 | 0.877 |
| Antipsychotics | -0.039 | 0.620 | 0.020 | 0.800 |

### F. Statistical Analysis – Breathing

**Table S12.** Breathing Main Summary. Mean feature per night distributions.

|  | ICU | Sleeplab All | S.Lab AHI<5 | S.Lab AHI>15 |
| --- | --- | --- | --- | --- |
| n subjects | 63 | 188 | 69 | 48 |
| Resp. Rate <sup>a)</sup> | 17.4 (3.2) | *** 15.9 (2.3) | *** 16.1 (2.2) | *** 15.7 (2.3) |
| Inter-Breath-Interval <sup>a)</sup> | 3.9 (0.8) | *** 4.7 (0.8) | *** 4.5 (0.7) | *** 5.0 (0.9) |
| Variability Index <sup>b)</sup> | 0.17 (0.14) | 0.18 (0.1) | 0.15 (0.07) | *** 0.2 (0.1) |
| Ventilation CVar <sup>b)</sup> | 0.16 (0.13) | * 0.2 (0.12) | 0.16 (0.09) | *** 0.3 (0.15) |
| <sup>a)</sup> mean (std), <sup>b)</sup> median (iqr) |  |  |  |  |

**Table S13.** Breathing feature analysis –feature mean (e.g. mean respiratory rate) per night, per patient.

|  | Sleep |  |  |  | Stage N1 |  |  |  | Stage N2 |  |  |  |
| --- | --- | --- | --- | --- | --- | --- | --- | --- | --- | --- | --- | --- |
|  | RR | IBI | Var | Vent | RR | IBI | Var | Vent | RR | IBI | Var | Vent |
| ICU, N_Subj | 63 | 63 | 63 | 63 | 56 | 56 | 56 | 56 | 38 | 38 | 38 | 38 |
| Lab All, N_Subj | 188 | 188 | 188 | 188 | 185 | 185 | 185 | 185 | 167 | 167 | 167 | 167 |
| Lab AHI 0-5, N_Subj | 69 | 69 | 69 | 69 | 66 | 66 | 66 | 66 | 64 | 64 | 64 | 64 |
| Lab AHI 15-100, N_Subj | 48 | 48 | 48 | 48 | 48 | 48 | 48 | 48 | 37 | 37 | 37 | 37 |
| ICU, Mean | 17.4 | 3.9 | 0.2 | 0.19 | 17.3 | 4 | 0.23 | 0.22 | 17.1 | 4 | 0.19 | 0.19 |
| ICU, Median | 17.4 | 3.8 | 0.17 | 0.16 | 17.3 | 3.9 | 0.21 | 0.2 | 17.6 | 3.7 | 0.14 | 0.13 |
| Lab All, Mean | ** 15.9 | 4.8 | 0.2 | 0.24 | ** 15.4 | ** 5.2 | ** 0.3 | 0.31 | * 15.9 | 4.6 | 0.18 | 0.21 |
| Lab All, Median | 15.8 | ** 4.7 | 0.18 | * 0.2 | 15.2 | 5.1 | 0.26 | ** 0.3 | 16 | ** 4.5 | 0.16 | 0.18 |
| Lab AHI 0-5, Mean | ** 16.1 | ** 4.5 | 0.17 | 0.19 | ** 15.6 | ** 5.0 | 0.23 | 0.27 | 16.1 | ** 4.4 | 0.16 | 0.18 |
| Lab AHI 0-5, Median | 16.2 | 4.5 | 0.15 | 0.17 | 15.3 | 4.9 | 0.22 | * 0.2 | 16.2 | 4.5 | 0.13 | 0.15 |
| Lab AHI 15-100, Mean | ** 15.7 | ** 5.0 | 0.27 | 0.32 | 15.2 | ** 5.4 | ** 0.3 | 0.38 | 15.8 | ** 4.8 | 0.24 | 0.29 |
| Lab AHI 15-100, Median | 15.8 | 4.9 | ** 0.2 | ** 0.3 | ** 15.2 | 5.2 | 0.3 | ** 0.4 | 15.7 | 4.6 | * 0.2 | * 0.3 |
| ICU, Std | 3.2 | 0.8 | 0.09 | 0.1 | 3.2 | 0.8 | 0.08 | 0.09 | 3.4 | 0.9 | 0.12 | 0.13 |
| ICU, Iqr | 4 | 1.1 | 0.14 | 0.13 | 3.6 | 0.9 | 0.13 | 0.12 | 5.5 | 1.4 | 0.14 | 0.15 |
| ICU, p_dag | 0.2637 | 0.1625 | 0.0297 | 0.0031 | 0.1351 | 0.2717 | 0.1091 | 0.0415 | 0.3823 | 0.1913 | 0.001 | 0.0001 |
| ICU, p_sha | 0.4523 | 0.0483 | 0.0013 | 0.0001 | 0.363 | 0.1206 | 0.0096 | 0.0025 | 0.0632 | 0.0254 | 0 | 0 |
| ICU, gaussian | True | True | False | False | True | True | True | False | True | True | False | False |
| Lab All, p_tt | 0 | 0 | 0.6084 | 0.0056 | 0 | 0 | 0.0025 | 0 | 0.0127 | 0 | 0.6373 | 0.2537 |
| Lab All, p_medians | 0.0032 | 0 | 0.799 | 0.0453 | 0.0001 | 0 | 0.0515 | 0.0002 | 0.0988 | 0.0025 | 0.2206 | 0.0519 |
| Lab All, p_mwu | 0.0001 | 0 | 0.1563 | 0.0006 | 0 | 0 | 0.0007 | 0 | 0.0242 | 0 | 0.1591 | 0.0085 |
| Lab All, Std | 2.3 | 0.8 | 0.08 | 0.11 | 2.1 | 0.9 | 0.08 | 0.12 | 2.4 | 0.8 | 0.08 | 0.1 |
| Lab All, Iqr | 3.1 | 1 | 0.1 | 0.12 | 2.7 | 1.1 | 0.12 | 0.15 | 3.4 | 0.9 | 0.09 | 0.11 |
| Lab All, p_dag | 0.3474 | 0.0004 | 0 | 0 | 0.4243 | 0.1074 | 0.2931 | 0 | 0.4012 | 0.0002 | 0 | 0 |
| Lab All, p_sha | 0.6601 | 0.0001 | 0 | 0 | 0.2886 | 0.0519 | 0.2004 | 0 | 0.5498 | 0.0004 | 0 | 0 |
| Lab All, gaussian | True | False | False | False | True | True | True | False | True | False | False | False |
| Lab AHI 0-5, p_tt | 0.0047 | 0 | 0.0832 | 0.9502 | 0.0006 | 0 | 0.6386 | 0.0065 | 0.0887 | 0.0033 | 0.1018 | 0.572 |
| Lab AHI 0-5, p_medians | 0.0814 | 0 | 0.2958 | 1 | 0.0064 | 0 | 0.5857 | 0.0182 | 0.3058 | 0.0078 | 0.8377 | 0.3058 |
| Lab AHI 0-5, p_mwu | 0.004 | 0 | 0.0776 | 0.3293 | 0.0006 | 0 | 0.2448 | 0.0035 | 0.0755 | 0.0015 | 0.2181 | 0.3103 |
| Lab AHI 0-5, Std | 2.2 | 0.7 | 0.07 | 0.09 | 2.1 | 0.9 | 0.08 | 0.11 | 2.2 | 0.7 | 0.07 | 0.09 |
| Lab AHI 0-5, Iqr | 2.9 | 0.9 | 0.07 | 0.09 | 3.3 | 1.1 | 0.13 | 0.16 | 3.2 | 0.7 | 0.07 | 0.09 |
| Lab AHI 0-5, p_dag | 0.8637 | 0.0214 | 0 | 0 | 0.2761 | 0.2972 | 0.0631 | 0.1523 | 0.4704 | 0.0214 | 0 | 0 |
| Lab AHI 0-5, p_sha | 0.9389 | 0.0885 | 0 | 0 | 0.2463 | 0.1272 | 0.1473 | 0.0609 | 0.2964 | 0.0615 | 0 | 0 |
| Lab AHI 0-5, gaussian | True | True | False | False | True | True | True | True | True | True | False | False |
| Lab AHI 15-100, p_tt | 0.0021 | 0 | 0.0001 | 0 | 0.0002 | 0 | 0 | 0 | 0.0973 | 0.0001 | 0.0392 | 0.0011 |
| Lab AHI 15-100, p_medians | 0.016 | 0 | 0.0002 | 0 | 0.0002 | 0 | 0.0008 | 0 | 0.0829 | 0.0039 | 0.0154 | 0.0154 |
| Lab AHI 15-100, p_mwu | 0.0009 | 0 | 0 | 0 | 0.0001 | 0 | 0 | 0 | 0.0388 | 0.0001 | 0.0009 | 0 |
| Lab AHI 15-100, Std | 2.3 | 0.9 | 0.08 | 0.12 | 2.2 | 0.9 | 0.08 | 0.13 | 2.9 | 0.9 | 0.09 | 0.13 |
| Lab AHI 15-100, Iqr | 2.5 | 1.5 | 0.1 | 0.13 | 2.3 | 1.4 | 0.11 | 0.17 | 3 | 1.5 | 0.11 | 0.14 |
| Lab AHI 15-100, p_dag | 0.6191 | 0.204 | 0.0778 | 0.0005 | 0.0283 | 0.462 | 0.2983 | 0.0056 | 0.0339 | 0.3913 | 0.1499 | 0.0009 |
| Lab AHI 15-100, p_sha | 0.8092 | 0.0346 | 0.0253 | 0.0009 | 0.0462 | 0.4451 | 0.1821 | 0.0092 | 0.1313 | 0.1887 | 0.0796 | 0.0053 |
| Lab AHI 15-100, gaussian | True | True | True | False | False | True | True | False | True | True | True | False |

|  | Stage N3 |  |  |  | Stage R |  |  |  | Wake |  |  |  |
| --- | --- | --- | --- | --- | --- | --- | --- | --- | --- | --- | --- | --- |
|  | RR | IBI | Var | Vent | RR | IBI | Var | Vent | RR | IBI | Var | Vent |
| ICU, N_Subj | 25 | 25 | 25 | 25 | 24 | 24 | 24 | 24 | 57 | 57 | 57 | 57 |
| Lab All, N_Subj | 79 | 79 | 79 | 79 | 124 | 124 | 124 | 124 | 180 | 180 | 180 | 180 |
| Lab AHI 0-5, N_Subj | 30 | 30 | 30 | 30 | 50 | 50 | 50 | 50 | 65 | 65 | 65 | 65 |
| Lab AHI 15-100, N_Subj | 16 | 16 | 16 | 16 | 24 | 24 | 24 | 24 | 48 | 48 | 48 | 48 |
| ICU, Mean | 18.8 | 3.5 | 0.13 | 0.12 | 19.1 | 3.5 | 0.19 | 0.16 | 17.9 | 3.9 | 0.23 | 0.21 |
| ICU, Median | 18.7 | 3.4 | 0.12 | 0.1 | 18.6 | 3.5 | 0.17 | 0.15 | 17.8 | 3.8 | 0.22 | 0.2 |
| Lab All, Mean | 16.7 | 4.3 | 0.1 | 0.1 | ** 16.4 | 4.4 | 0.2 | 0.22 | 15.7 | 5.3 | ** 0.3 | 0.31 |
| Lab All, Median | * 16.2 | ** 4.2 | 0.1 | 0.1 | 16.4 | ** 4.3 | 0.18 | * 0.2 | ** 15.6 | ** 5.3 | 0.28 | ** 0.3 |
| Lab AHI 0-5, Mean | 16.5 | 4.1 | 0.09 | 0.09 | ** 16.8 | ** 4.2 | 0.19 | * 0.2 | 15.8 | 5.2 | 0.25 | ** 0.3 |
| Lab AHI 0-5, Median | 16.6 | ** 4.1 | 0.09 | 0.09 | 16.7 | 4.1 | 0.17 | 0.18 | ** 15.6 | ** 5.1 | 0.25 | 0.28 |
| Lab AHI 15-100, Mean | 18.6 | 4 | 0.13 | 0.13 | ** 16.3 | ** 4.5 | 0.21 | ** 0.2 | 15.9 | 5.4 | ** 0.3 | ** 0.3 |
| Lab AHI 15-100, Median | 18.2 | 4 | 0.12 | 0.12 | 16 | 4.4 | 0.19 | 0.21 | ** 15.9 | ** 5.3 | 0.29 | 0.34 |
| ICU, Std | 4.2 | 0.8 | 0.06 | 0.06 | 2.9 | 0.6 | 0.06 | 0.06 | 3 | 0.7 | 0.06 | 0.06 |
| ICU, Iqr | 3.1 | 0.6 | 0.05 | 0.04 | 4 | 0.7 | 0.06 | 0.06 | 2.6 | 0.6 | 0.1 | 0.09 |
| ICU, p_dag | 0.0467 | 0.0008 | 0 | 0 | 0.6781 | 0.0469 | 0.3604 | 0.1157 | 0.0054 | 0.0428 | 0.1557 | 0.1384 |
| ICU, p_sha | 0.2026 | 0.007 | 0.0002 | 0 | 0.2319 | 0.0784 | 0.0745 | 0.06 | 0.0053 | 0.0165 | 0.0506 | 0.0222 |
| ICU, gaussian | True | False | False | False | True | True | True | True | False | False | True | True |
| Lab All, p_tt | 0.0192 | 0.0002 | 0.0219 | 0.0602 | 0 | 0 | 0.6089 | 0.0067 | 0 | 0 | 0.0001 | 0 |
| Lab All, p_medians | 0.0217 | 0.0013 | 0.1686 | 0.3587 | 0.0001 | 0 | 0.8235 | 0.0142 | 0 | 0 | 0.007 | 0 |
| Lab All, p_mwu | 0.0046 | 0 | 0.0079 | 0.0351 | 0 | 0 | 0.4493 | 0.0012 | 0 | 0 | 0 | 0 |
| Lab All, Std | 3.7 | 0.9 | 0.04 | 0.04 | 2.4 | 0.8 | 0.07 | 0.1 | 2 | 0.9 | 0.07 | 0.11 |
| Lab All, Iqr | 4.4 | 1 | 0.04 | 0.04 | 3.3 | 0.9 | 0.08 | 0.12 | 2.8 | 1.2 | 0.1 | 0.14 |
| Lab All, p_dag | 0.0007 | 0.0158 | 0 | 0 | 0.8617 | 0.0007 | 0 | 0 | 0.8296 | 0.0284 | 0.4131 | 0.0045 |
| Lab All, p_sha | 0.0012 | 0.0587 | 0 | 0 | 0.9268 | 0.0002 | 0 | 0 | 0.9074 | 0.0042 | 0.3842 | 0.0011 |
| Lab All, gaussian | False | True | False | False | True | False | False | False | True | False | True | False |
| Lab AHI 0-5, p_tt | 0.0277 | 0.004 | 0.0017 | 0.0154 | 0.0002 | 0 | 0.8247 | 0.0314 | 0 | 0 | 0.0897 | 0 |
| Lab AHI 0-5, p_medians | 0.0804 | 0.0018 | 0.0804 | 0.0804 | 0.0063 | 0 | 1 | 0.0822 | 0 | 0 | 0.1466 | 0.0003 |
| Lab AHI 0-5, p_mwu | 0.0045 | 0.0004 | 0.001 | 0.0032 | 0.0002 | 0 | 0.3409 | 0.0148 | 0 | 0 | 0.0467 | 0 |
| Lab AHI 0-5, Std | 3.2 | 0.7 | 0.02 | 0.02 | 2.1 | 0.7 | 0.06 | 0.07 | 2.2 | 1 | 0.08 | 0.11 |
| Lab AHI 0-5, Iqr | 3.5 | 0.8 | 0.03 | 0.03 | 2.7 | 0.8 | 0.08 | 0.11 | 2.8 | 1.3 | 0.1 | 0.15 |
| Lab AHI 0-5, p_dag | 0 | 0.8084 | 0.6682 | 0.6959 | 0.8907 | 0.0789 | 0.2226 | 0.1576 | 0.8125 | 0.0024 | 0.2555 | 0.2669 |
| Lab AHI 0-5, p_sha | 0.0028 | 0.9955 | 0.7991 | 0.3258 | 0.8184 | 0.291 | 0.0405 | 0.0443 | 0.939 | 0.0004 | 0.1907 | 0.0755 |
| Lab AHI 0-5, gaussian | False | True | True | True | True | True | True | True | True | False | True | True |
| Lab AHI 15-100, p_tt | 0.8935 | 0.09 | 0.8368 | 0.8155 | 0.001 | 0 | 0.356 | 0.0078 | 0.0001 | 0 | 0 | 0 |
| Lab AHI 15-100, p_medians | 0.8452 | 0.0843 | 0.8452 | 0.2776 | 0.0015 | 0.0002 | 0.7728 | 0.1489 | 0.0012 | 0 | 0.0001 | 0 |
| Lab AHI 15-100, p_mwu | 0.4521 | 0.0263 | 0.2919 | 0.1172 | 0.0004 | 0 | 0.2514 | 0.0045 | 0 | 0 | 0 | 0 |
| Lab AHI 15-100, Std | 4.4 | 1 | 0.04 | 0.03 | 2.7 | 0.9 | 0.08 | 0.1 | 2 | 0.8 | 0.07 | 0.1 |
| Lab AHI 15-100, Iqr | 6 | 1 | 0.08 | 0.05 | 2.3 | 1.2 | 0.12 | 0.14 | 2.8 | 1.3 | 0.09 | 0.13 |
| Lab AHI 15-100, p_dag | 0.5921 | 0.2752 | 0.3055 | 0.6715 | 0.2161 | 0.5336 | 0.1563 | 0.0321 | 0.903 | 0.3198 | 0.99 | 0.4688 |
| Lab AHI 15-100, p_sha | 0.7358 | 0.7662 | 0.2263 | 0.8064 | 0.1637 | 0.3835 | 0.11 | 0.057 | 0.7408 | 0.2948 | 0.9796 | 0.4818 |
| Lab AHI 15-100, gaussian | True | True | True | True | True | True | True | True | True | True | True | True |

**Figure S6.**
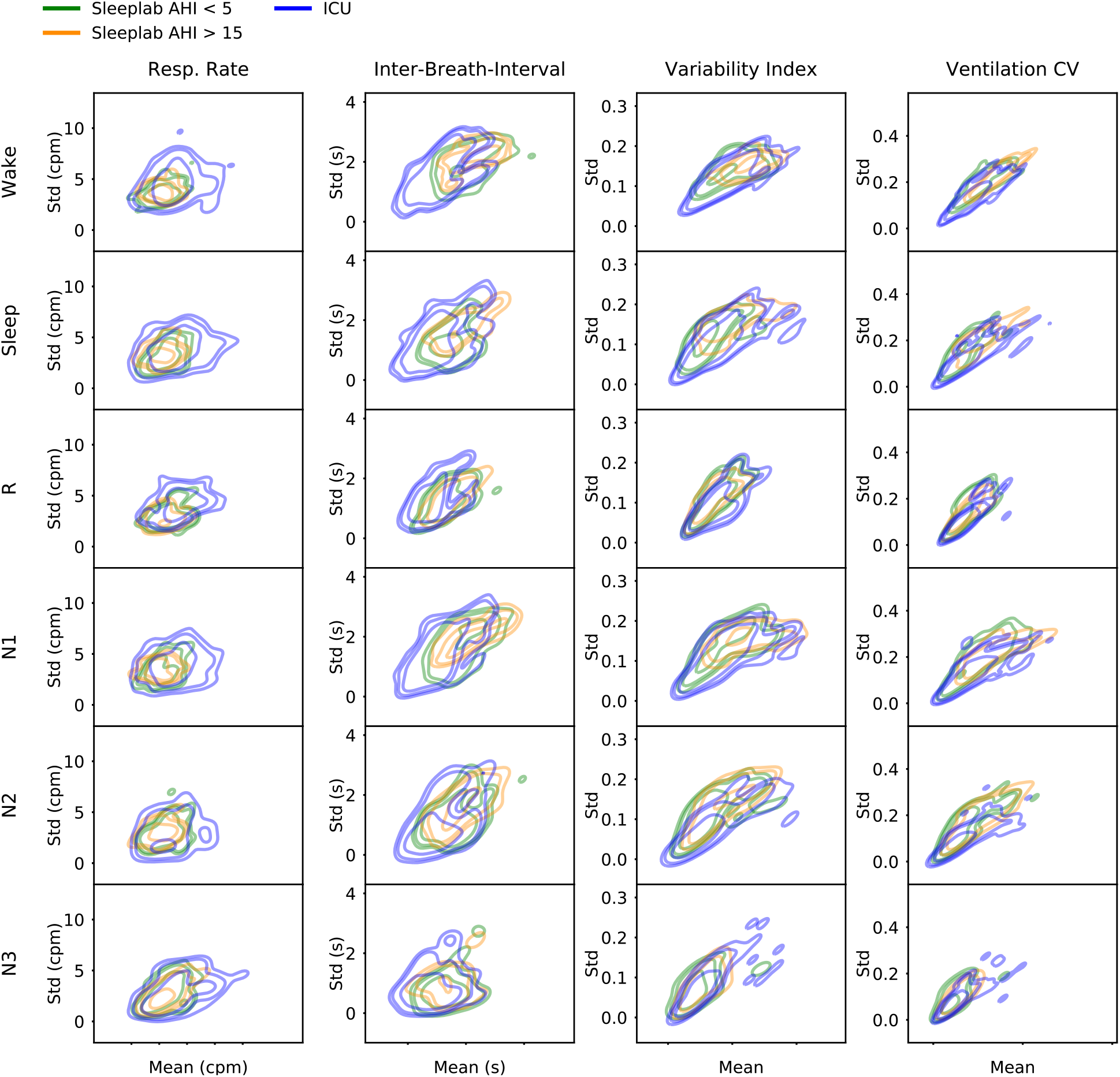
Breathing features per night (mean and standard deviation) for each sleep stage and for ICU and sleeplab AHI<5 and AHI>15 cohort. Distributions were fit with a kernel density estimation method, iso mass levels 0.1, 0.2, and 0.5 are shown in the plot.

## Notes

### Funding Statement

M.B.W was supported by the Glenn Foundation for Medical Research and American Federation for Aging Research (Breakthroughs in Gerontology Grant); American Academy of Sleep Medicine (AASM Foundation Strategic Research Award); Football Players Health Study (FPHS) at Harvard University; Department of Defense through a subcontract from Moberg ICU Solutions, Inc; and NIH (1R01NS102190, 1R01NS102574, 1R01NS107291, 1RF1AG064312).

### Author Declarations

The study was approved by the Mass General Brigham Institutional Review Board.

